# Mathematical modeling suggests improved clinical outcomes of second-generation PARP inhibitors with reduced toxicity

**DOI:** 10.1101/2024.01.29.24301918

**Authors:** Emilia Kozłowska, Ulla-Maija Haltia, Krzysztof Puszynski, Anniina Färkkilä

## Abstract

High-grade serous ovarian cancer (HGSC) is one of the most lethal gynecological cancers. Recent clinical trials have shown the remarkable benefits of maintenance treatment with Poly-ADP Ribose Polymerase inhibitors, particularly in patients with tumors deficient in homologous recombination DNA repair. However, the large Phase III clinical trials are limited in their ability to evaluate various dosages and treatment durations, and the effects of toxicities and treatment interruptions on clinical outcomes remain unknown. In this study, we developed a computational framework for virtual clinical trials taking into account both tumor dynamics (growth selection and evolution of resistance mechanisms) and clinico-pharmacological features (pharmacokinetics, dosing, hematological toxicity, dose interruptions, and reductions). Our model replicates the clinical outcomes observed in the landmark SOLO-1 clinical trial. The branching process model revealed a heterogeneous patient population with distinct tumor dynamics, highlighting fitness and selection pressure as determinants of a poor response. Through a virtual trial approach, we show that managing toxicity via treatment interruptions or dose reductions does not compromise the clinical benefits of the treatment. Importantly, we present evidence that further reduction of hematological toxicity could significantly improve clinical outcomes in first-line PARPi maintenance treatment in ovarian cancer.

**Significance:** Phase III clinical trials are limited in their capacity to investigate different dosing schedules, and the number of participants is relatively small. Furthermore, the effects of toxicities on trial outcomes are frequently unknown. Virtual clinical trials using a mechanistic mathematical model combined with statistical evaluations can overcome these limitations. In this work, we demonstrate the application of a computational framework for optimizing drug scheduling and testing of the next generation of PARP inhibitors, providing insights to guide future clinical trial design.

## Introduction

High-grade serous ovarian cancer (HGSC) is the most common and lethal subtype of ovarian cancer, typically diagnosed at an advanced stage (1,2). The cornerstone of therapy for newly diagnosed HGSC is cytoreductive surgery combined with platinum-based chemotherapy (3). However, nearly all patients experience relapse with a tumor that develops resistance to conventional chemotherapeutic approaches (4–6), resulting in poor overall survival (7).

Over the past decade, Poly-ADP Ribose Polymerase (PARP) inhibitors have revolutionized HGSC treatment, particularly in patients with germline or somatic *BRCA1/2* mutations or tumors with homologous recombination deficiency (HRD) (8,9). PARP inhibitors are a group of pharmacological inhibitors of PARP enzymes, which play roles in transcription regulation, apoptosis, and DNA damage response. In HRD cells, the inhibition of PARP prevents the DNA repair process, disrupting cellular homeostasis and ultimately leading to cell death (10). Approximately 50% of ovarian cancer patients with HRD and/or sensitivity to platinum-based chemotherapy are especially sensitive to PARP inhibitors (11). Thus, HRD and platinum sensitivity are the primary biomarkers for PARPi response (11,12)

Olaparib was among the first PARPi that had favorable outcomes in the SOLO-1 clinical trial as a first-line maintenance treatment after debulking surgery and platinum-based chemotherapy in patients with germline *BRCA1/2* mutations (13). Currently, three PARP inhibitors have been approved by the U.S. Food and Drug Administration (FDA) and European Medicines Agency (EMA) for the treatment of ovarian cancer: olaparib (in 2014) (14), rucaparib (in 2016) (15), and niraparib (in 2017) (16).

Despite recommendations for schedules and doses based on results from phase I/II–III clinical trials, not all potential treatment protocols can be clinically tested due to practical and ethical constraints. Therefore, different dosing schemes, treatment durations, and the effects of toxicities and treatment interruptions on clinical outcomes remain unknown. In addition, not all patients respond to PARPi maintenance treatment, and acquired resistance to PARP inhibitors is an emerging clinical problem. Previous preclinical and translational work has suggested that acquired PARPi resistance is associated with significant clonal, genomic, and functional heterogeneity (17,18). However, because clinical biopsies are not typically performed upon PARPi resistance after first-line maintenance, the clonal dynamics and their clinical relevance are unknown. Thus, virtual trials based on mathematical modeling can provide valuable supporting evidence for the clinical testing of various hypotheses regarding novel therapeutic agents (19,20).

In this study, we employed a computational approach to optimize the maintenance treatment with PARPi in high-grade serous ovarian cancer. By calibrating the mathematical model on the SOLO-1 clinical trial, we investigated the effects of treatment duration, optimal dosing, as well as the management of hematological toxicity. We showed that our model faithfully reproduces real-world clinical outcomes and applied the fitted model to investigate whether the clonal dynamics, treatment adjustments, and toxicity management affect the clinical outcomes. Importantly, we present evidence that further reduction of hematological toxicity could lead to significantly improved clinical outcomes upon first-line PARPi maintenance treatment in ovarian cancer.

## Materials and Methods

### SOLO-1 clinical trial data

SOLO-1 is a Phase III randomized, double-blinded, placebo-controlled clinical trial designed to evaluate the efficacy and safety of olaparib tablets (300 mg twice a day) as a maintenance monotherapy compared with a placebo. The admission criteria for the trial were as follows: 1) newly diagnosed advanced *BRCA1/2* mutated ovarian cancer, 2) the patient must be in complete or partial response following platinum-based chemotherapy, and 3) the patient must have normal bone marrow function. In total, 391 patients were enrolled in this trial. Patients were randomized to receive olaparib or a placebo for 2 years or, in the case of relapse, until disease progression. The primary endpoint of the trial was progression-free survival, and the toxicity profile was also assessed.

In our study, the mathematical model was fitted to data on progression-free survival rates (depicted as a Kaplan–Meier plot) from the SOLO-1 clinical trial. In addition, the hematological toxicity profiles of Olaparib (from the SOLO-1 trial) and platinum-based chemotherapy (21) were extracted to fit the model parameters to include hematological toxicity in the model.

### Stochastic mathematical model

We developed a mechanistic model consisting of three layers. The first layer is the cancer cell dynamics. The second is the pharmacokinetics of the drugs, and the last one is the hematological toxicity of the drug measured by the number of white blood cells (WBCs). We included cancer cells as a heterogeneous, well-mixed mass differing in the level of resistance to carboplatin and olaparib. During tumor growth, both with and without treatment, the accumulation of (epi)genetic aberrations results in cancer cells gaining new resistance mechanisms. Each cell is defined by two numbers, where the first and second ones denote the number of resistance mechanisms to carboplatin and olaparib, respectively. For example, a cell defined as “1,3” has one and three resistance mechanisms to carboplatin and olaparib, respectively. The number of resistance mechanisms that could accumulate in cancer cells is not restricted and can go to infinity. The only limitation is the tumor burden reaching a level where symptoms become clinically evident, and the patient relapses.

The second layer, describing the pharmacokinetics of carboplatin and olaparib, is included as a one-compartmental model depicting exponential drug decay. Because the time interval from drug administration to the highest drug concentration in the peripheral blood (*T*_*max*_), is in hours, and simulations occur on a daily scale, we assume that *T*_*max*_=0. In such a way, we modeled intravenous bolus intravenous or peroral administration for carboplatin cisplatin and olaparib, respectively.

Subsequently, we included the hematological toxicity of platinum-based chemotherapy and olaparib in the model. Because hematological toxicity is associated with a low level of WBCs, we modeled the dynamics of WBCs throughout treatment and treatment intervals. In the absence of treatment, the number of WBCs is at dynamic equilibrium. During treatment, however, the level of WBCs decreases. We modeled WBC growth as a density-limiting growth using a logistic model. This allows WBCs to oscillate around the equilibrium in the absence of treatment and the decay of WBC during treatment intervention, as explained in detail in Supplementary Text 5.

### A computer simulator of HGSC patient course

We developed a simulator for the course of HGSC patients from cancer initiation to the second recurrence. The computer simulations include the following phases: 1) pre-diagnosis, 2) primary treatment phase, 3) maintenance treatment phase, 4) post-treatment phase, 5) second-line treatment, and 6) the second post-treatment phase.

During the pre-diagnosis phase, the tumor grows from a single sensitive cell (“0,0”) without any perturbations. During tumor growth, there is a probability that a cell will gain one resistance mechanism to one of two drugs during cell division. During this simulation phase, the tumor evolves according to the neutral evolution law. This phase concludes when the tumor reaches *M* cancer cells, which reflects the time of diagnosis. We assume that treatment starts at the time of diagnosis without any loss of generality.

The primary treatment phase starts with upfront cytoreductive surgery (primary debulking surgery - PDS). Surgery is modeled by the removal of a fraction of cancer cells at a one-time point, followed by a 3-week recovery period and platinum-based chemotherapy. Chemotherapy is administered as an intravenous injection (bolus IV). Each patient in the virtual cohort undergoes 6 cycles of platinum-based chemotherapy, with the median interval between them equal to 3 weeks. Maintenance treatment with olaparib starts after the WBC level is within the normal range after chemotherapy. Patients then receive olaparib as peroral administration twice a day. This phase is simulated for 2 years, until relapse or if toxicity becomes too high, depending on which time is shorter. Subsequently, the patient is simulated in a follow-up until relapse, if relapse does not occur during maintenance treatment. During this period, the tumor grows without perturbation as the patient is asymptomatic, and the tumor remains undetectable. The phase concludes with the first relapse. The subsequent phase is the second-line platinum-based chemotherapy. Similar to the primary treatment, patients receive 6 cycles of chemotherapy at a 3-week interval. The final phase of the simulation is a follow-up after the second-line treatment. Here, cancer cells can grow without perturbation, reflecting the time when the patient is in clinical remission or receives palliative treatment, depending on the chemotherapy response. This phase concludes with the second relapse. The main output of the simulation is a response rate after the first- and second-line treatments, measured as progression-free survival and toxicity level to platinum-based chemotherapy and olaparib. The simulator is freely available on GitHub at https://github.com/EmiliaKo/VCT_olaparib.

### Mathematical model calibration using SOLO-1 clinical trial and toxicity profile data

The developed mathematical model was fitted to two main types of data: SOLO-1 clinical trial data (comprising treatment protocol and response rate in the form of progression-free survival) and the toxicity profile of carboplatin and PARPi. A detailed description of model calibration and a list of all parameters along with their values are provided in Supplementary Text 7 and Supplementary Table S4, respectively. In brief, we initially set all model parameters that could be directly extracted from clinical data as constant values. Subsequently, we fitted toxicity parameters to align with the toxicity profile of the two drugs included in the model (carboplatin and olaparib). In the final step, we fitted the remaining parameters to ensure that the Kaplan– Meier plot produced from the model simulations agrees with the one obtained from the SOLO-1 clinical trial. A detailed description of the model fitting process is presented in Supplementary Text 7.

### Tumor feature extraction and Cox regression

We extracted the outputs of the model simulations that describe tumor dynamics and heterogeneity. Subsequently, we performed Cox regression on a cohort of 10,000 virtual patients, utilizing all extracted features (see Supplementary Text 10 and Supplementary Table 6). As a metric of Cox regression performance, we applied the concordance index (c-index). We performed the analysis only in the univariate scenario using 10-fold cross-validation.

### Treatment duration optimization

In the optimization of treatment duration, we simulated 10,000 virtual HGSC patients under a given condition and calculated the median PFS. For maintenance duration, we initially conducted a simulation without any maintenance treatment to establish a baseline. Subsequently, we incrementally increased the duration of olaparib maintenance by 1 month at a time. We assessed the impact of the extended duration on PFS and continued extending the treatment period in the event of a significant increase in PFS until no further difference was observed. Similarly, we optimized the olaparib dose.

### Toxicity management

We also investigated the performance of a new generation of PARPi that is less toxic by reducing the parameter responsible for decreasing WBC level during olaparib administration. The toxicity level was assessed by calculating the percentage of virtual patients who developed hematological toxicity to olaparib. Importantly, we incorporated three methods for dealing with olaparib toxicity: dose reduction, dose interruption, and treatment discontinuation. From real-world data, we estimated that a total of 40% of the patients had grade 2–3 hematological toxicity, resulting in one or several forms of toxicity management in SOLO-1 (22). However, real-world data were not available on the proportions of these patients who underwent more than one toxicity management procedure. Thus, in the model, upon hematological toxicity, each patient would undergo only one toxicity management, and after repeated toxicity, the treatment was discontinued. A detailed description of the methods to overcome olaparib toxicity is provided in Supplementary Text 8.

## Results

### An integrative computational framework for virtual clinical trials of 1^st^-line maintenance treatment of cancer patients

We developed an integrated computational framework for conducting clinical trials on cancer patients, considering the following treatment modalities: surgery, chemotherapy, and maintenance treatment (Figure 1). The platform includes a mechanistic model of cancer cell dynamics that is simulated numerically and a statistical model represented by a Kaplan–Meier estimator.

**Figure 1.**
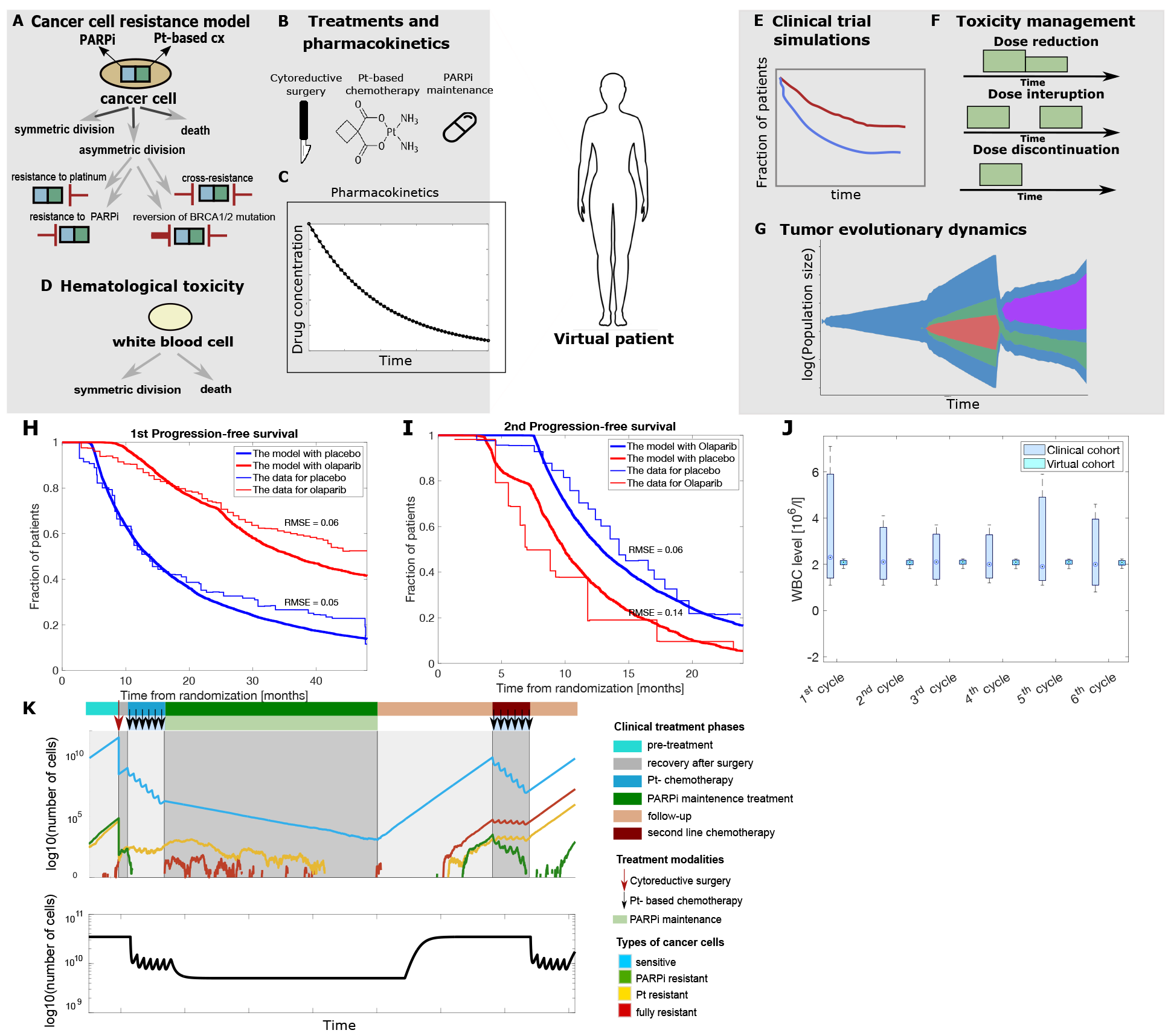
Mathematical modeling framework for virtual clinical trials of 1^st^-line maintenance treatment of ovarian cancer patients. A) Cancer cells in the model differ in the level of drug resistance to platinum-based chemotherapy (Pt-based CX) and targeted treatment with PARPi (olaparib). Resistance mechanisms can exist in a cancer cell either at diagnosis or develop during cancer treatment, with variably resistant clones formed through asymmetric division. B) The model includes three types of treatment: surgery, chemotherapy (with platinum compounds), and targeted treatment with PARPi (olaparib). C) Pharmacokinetics of chemotherapy and targeted treatment are modeled using a one-compartmental model. D) To evaluate hematological toxicity, white blood cell count is included as a one-type branching process model. E) The developed model was applied to simulate the SOLO-1 clinical trial by integrating the mathematical model with the Kaplan–Meier estimator. F) We utilized the model to assess the most suitable method for managing adverse events with olaparib. G) Tumor evolutionary dynamics of drug resistance to chemotherapy and PARPi were extracted to reveal features associated with response and resistance. H) The first progression-free survival, and I) the second progression-free survival from the SOLO-1 clinical trial and the model simulations. To evaluate the goodness of fit between our model and real-life PFS plots, RMSE was used as a statistical method. Our model shows an excellent fit to the real-life data. J) The lowest value of white blood cells (WBCs) after each chemotherapy cycle (from clinical and virtual patient cohorts). K) The tumor burden and the level of WBCs over time in a single exemplary virtual patient.

The mechanistic model is a modified branching process that is a discrete-time and discrete-state model, including a discrete-time model of olaparib and platinum-based chemotherapy pharmacokinetics and WBC dynamics with a discrete-time logistic growth model. In the model, the tumor is initiated from a single malignant cell and normal WBC levels. Cancer cells divide and mutate, resulting in a heterogeneous, well-mixed mass of cancer cells that can acquire resistance to chemotherapy (platinum) and PARP inhibitor both during the initial progression or during the treatments via the accumulation of (epi)genetic aberrations (23) (Figure 1A). The cancer treatment is modeled with debulking surgery and 1st-line chemotherapy, as described in Kozłowska et al. (2021), followed by maintenance treatment with a PARP inhibitor (Figure 1B). The pharmacokinetics of platinum-based chemotherapy and olaparib are represented by a one-compartmental model describing exponential drug elimination (Figure 1C). Because the time interval from drug administration to the highest drug concentration in the peripheral blood (Tmax), is in hours, and simulations are on a day scale, we assume that *T*_*max*_=0. Thus, we modeled intravenous bolus intravenous or peroral administration for carboplatin and olaparib, respectively. In the third layer, we included the hematological toxicity of platinum-based chemotherapy and olaparib, measured as the number of WBCs in the peripheral blood (Figure 1D), as an indicator of myelosuppression, including thrombocytopenia and anemia, which are common side effects of chemotherapy (21) and PARP inhibitors (22,24,25). Thus, we modeled the dynamics of WBCs using a discrete-time logistic model.

The mathematical model developed herein serves as the fundamental basis for conducting virtual clinical trials in cancer using the computational framework composed of a mathematical model and a Kaplan–Meier estimator as the output for clinical outcomes (Figure 1E). The framework can be applied to optimize the treatment (Figure 1F) and explore tumor evolutionary dynamics during PARPi maintenance treatment (Figure 1G). Notably, the developed model can be fitted and applied to other similar cancer types that follow the same treatment paradigm.

### Mathematical modeling framework faithfully reproduces the clinical outcomes of the SOLO-1 trial

First, we calibrated the model to match the clinical outcomes and toxicities observed in the landmark SOLO-1 clinical trial. The effects of surgery and chemotherapy on the cancer cells were adjusted as described previously (26). The toxicity parameter during chemotherapy was adjusted to the lowest level of WBCs after each cycle and the time when the lowest WBC value was observed (nadir), as adapted from a previous study (21). Olaparib toxicity was adjusted according to the reported rate of 40% of patients experiencing grade 3 hematologic toxicity that led to treatment interruption or dose reduction in the SOLO-1 trial (REF).

Our model showed an excellent fit to PFS (Figure 1H) of the SOLO-1 clinical trial, with a low root mean squared error (RMSE) serving as a metric for model fitness (0.06). Interestingly, real-world data suggest that the responses to platinum-based therapies in the second line have been remarkably poor in previously PARPi-treated patients (27). We then explored how the second PFS to platinum-based therapies of the model fit real-world data. Our observations indicated that our model reproduces the inferior responses to chemotherapy after PARPi maintenance observed in real-world data (Figure 1I). In addition, the WBC levels in the model matched the real-world data during the chemotherapy cycles (Figure 1J). Consequently, we successfully reproduced the clinical course, hematological toxicity, and tumor dynamics of virtual patients undergoing 1st-line treatment with surgery and platinum-based chemotherapy followed by PARPi maintenance (Figure 1K). Thus, the herein-developed mathematical framework can be used to statistically evaluate different treatment schemes and tumor dynamics to optimize the treatment.

### Cancer cell dynamics and heterogeneous patient populations suggest fitness, selection advantage, and chemosensitivity as determinants of PARPi response

To investigate cancer cell dynamics in a large patient population, we simulated 10,000 virtual ovarian cancer patients undergoing primary debulking surgery and adjuvant chemotherapy followed by PARP inhibitor maintenance therapy. Interestingly, we observed four distinct patterns of tumor dynamics in the patient groups (Figure 2A–E). The first and largest patient group (Figure 2A, “eradicated”) is characterized by a rapid decrease in sensitive and resistant cell populations during cytoreductive surgery and chemotherapy. In this patient population, only sensitive cells remained at the beginning of PARPi treatment, ultimately leading to the complete eradication of cancer cells. The second-largest patient population (Figure 2B, “heterogeneous”) is represented by partial responses to 1st-line treatment and rapid progression of the tumor during the PARPi maintenance treatment, resulting in a heterogeneous tumor with fully or partially resistant cells at relapse. The third-largest population (Figure 2C, “fully resistant”) was characterized by a fast emergence of a fully resistant population right after finishing chemotherapy. Moreover, we observed a relative overgrowth of resistant cells during PARPi treatment, but still, the presence of a sizeable fraction of sensitive cells at relapse after 2 years of PARPi maintenance treatment. The fourth patient population (Figure 2D, “sensitive”) is characterized by a slow progression of sensitive cells leading to relapse after a relatively long PFS>4 years. Even though there is a minority of resistant cells at relapse, these patients, at least partially, would likely respond to a second-line platinum-based chemotherapy. Interestingly, the Kaplan–Meier plot of the different patient populations revealed distinct responses to PARPi maintenance treatment among the patient groups (Figure 2F). As expected, the “eradicated”, and “sensitive” had an extremely favorable prognosis, whereas the “fully resistant” had an intermediate ____, and the “heterogeneous” patient group had the shortest PFS to 1st-line olaparib maintenance. For the fully resistant patients, this can be potentially explained by the initial presence of sensitive cancer cells and their gradual replacement by the fully resistant cells at relapse, whereas different partially resistant cell populations arise earlier in the “heterogeneous” patient group.

**Figure 2.**
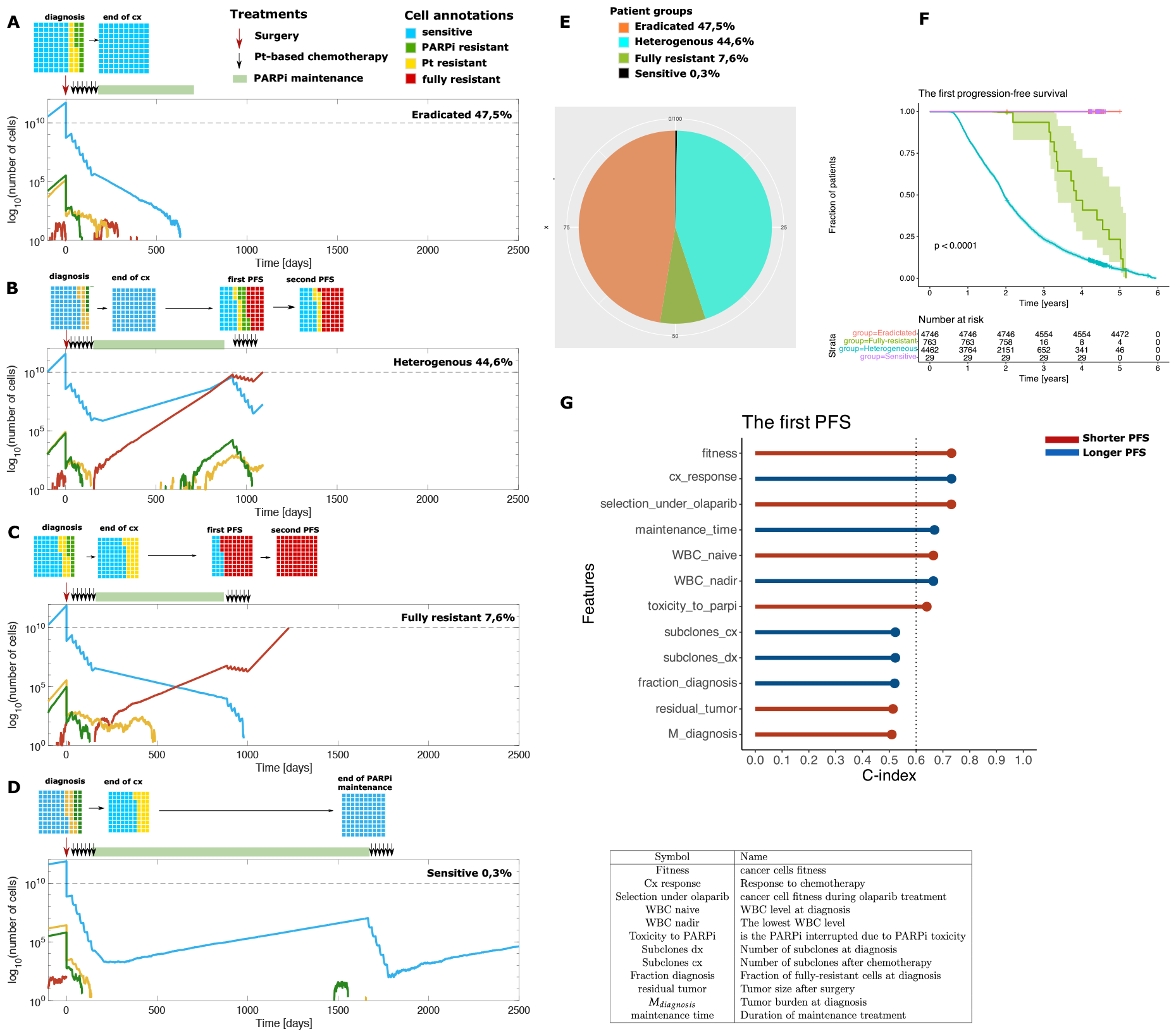
PARPi maintenance treatment induces heterogeneous cancer cell dynamics and patient populations with distinct clinical outcomes. A) eradicated, B) heterogenous, C) fully resistant, and D) sensitive. E) Proportions of each distinct clinical outcome in a virtual patient cohort. F) Kaplan–Meier estimator for the first progression-free survival of each of the four patient types. G) Model parameters associated with the response and resistance to PARPi. Univariate Cox regression for features related to tumor dynamics, with the c-index as a metric of model performance. Features within the rectangle show good predictive performance.

To further investigate which model features are most significantly associated with PFS on PARP inhibitors, we extracted the outputs of the model simulations that describe tumor growth and heterogeneity (Figure 2G) and applied the simulations with each feature, followed by the Cox regression model with 10-fold cross-validation. Interestingly, seven parameters were significantly associated with PFS, indicating that PFS is multifactorial, and tumor intrinsic resistance mechanisms do not define PFS as a single feature. The fitness of the cancer cells (i.e., selection advantage, which defines how fast the cancer cells are growing in comparison with other cells in the tumor) and selection under olaparib were the two most significant features associated with a shorter PFS. In addition, the WBC levels at diagnosis, time of maintenance treatment, and discontinuation of PARPi treatment due to toxicity were also associated with shorter PFS. By contrast, the parameters significantly associated with prolonged PFS were the response to first-line chemotherapy (measured as a fraction of cancer cells killed by chemotherapy), and reduced lowering of the white blood cells as an indicator of lower hematological toxicity.

### Drug dosing and toxicity management affect clinical outcomes of PARPi maintenance treatment

In the design of clinical trials, the maximum tolerated dose of the drug from phase I–II is typically utilized in phase III to investigate the efficacy of clinical outcomes. However, the impact of different drug doses and dose reductions on clinical outcomes is not well understood. Therefore, we set out to investigate the effect of different starting doses of olaparib as maintenance treatment using our integrative computational platform. We performed a simulation of 10,000 virtual HGSC patients treated with a wide range of olaparib doses between 100 mg to 400 mg twice a day, and the median PFS was computed (Figure 3A). We observed significant increases in PFS times with olaparib starting dose increases between 200 to 250 and 250 to 300 mg and a drop in doses higher than 300 mg twice a day (Figure 3A), suggesting that the toxicities included in the model can result in inferior clinical outcomes with doses over 300 mg bi-daily. Next, we investigated the optimal duration of maintenance treatment and observed, revealing that the optimal treatment duration with olaparib was 27 months (Figure 3B). Notably, no additional benefit was obtained from prolonged olaparib maintenance beyond 27 months. We then examined the dose reductions from the starting dose of 300 mg twice a day after toxicity and noted that no differences were observed in PFS, indicating that dose reduction due to hematological toxicity, even down to a 50 mg bi-daily dose, does not impair the efficacy of PARPi maintenance (Figure 3C). To investigate the effect of treatment interruptions, we included treatment breaks from 1 to 10 weeks in the model. In our analyses, the interruption of treatment in that range does not decrease or increase the first PFS (Figure 3D).

**Figure 3.**
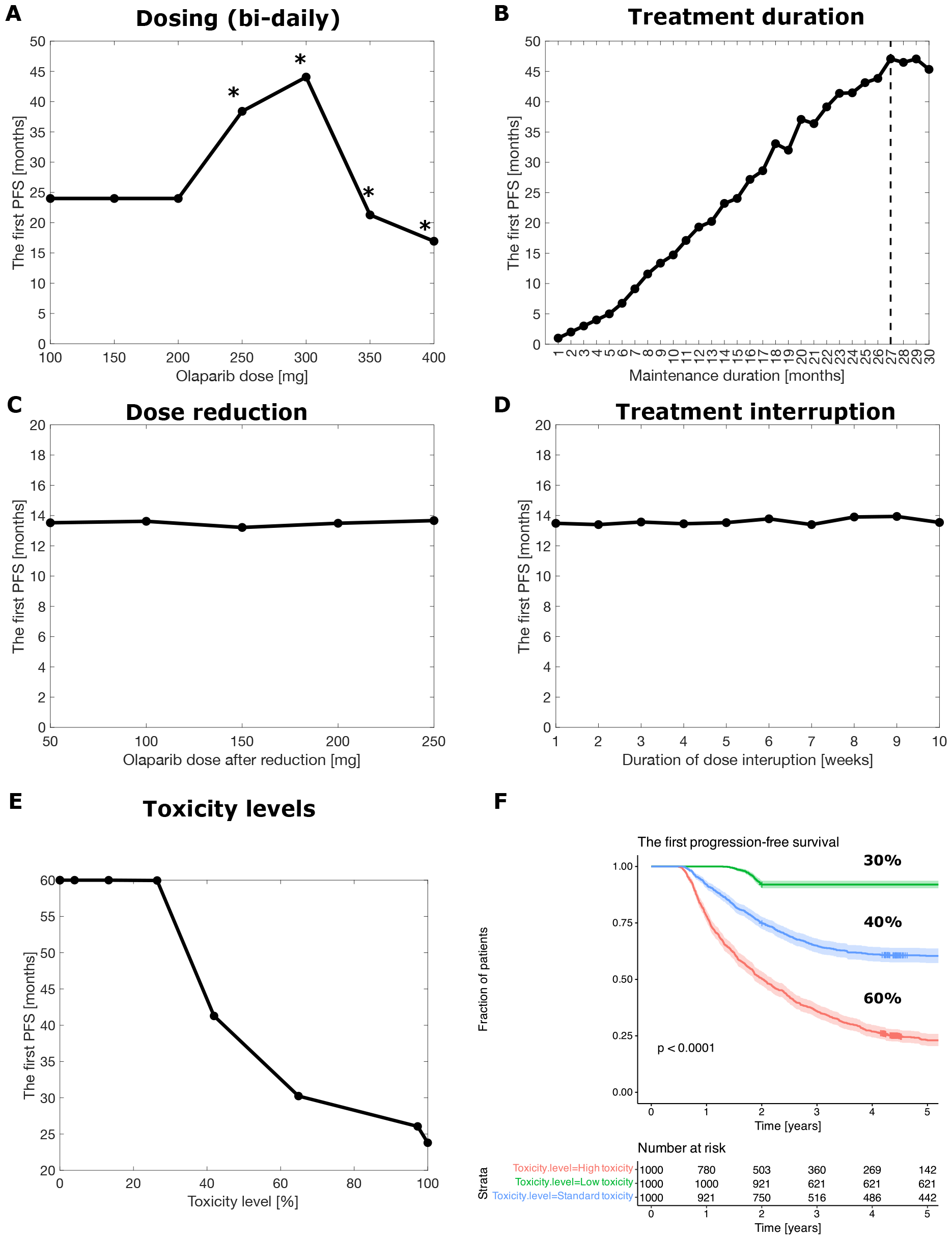
Effects of PARPi dosing, duration, and toxicity management on PFS. A) The first PFS as a function of A) olaparib dose and B) treatment duration. C) Effect of dose reduction on the first PFS. D) Effect of the duration of treatment interruption on the first PFS. E) The first PFS, as a function of the toxicity level, measured as a percentage of patients requiring toxicity management. F) The Kaplan–Meier estimator for patients with higher-than-normal levels of PARPi toxicity (60%), olaparib (40%), and the second-generation PARPi (30%). Asterisks indicate statistical significance compared to the previous dose with p<0.05 (t-test).

Recently, more specific 2nd generation PARP inhibitors have been introduced with lower toxicity profiles and potentially better-tolerated selective targeting of the PARP-1 enzyme (28). Lastly, we investigated how the administration of PARPis with similar potency but different toxicity levels affects PFS. We observed a significant shortening of PFS with toxicity levels higher than 40% (Figure 3E). Importantly, the reduction of toxicity to a level where 30% of patients undergo at least one form of toxicity management, such as the recently developed 2nd generation PARP1 specific inhibitors, led to a significantly prolonged PFS (log-rank test p-value<0.0001) (Figure 3F). Thus, our results suggest significant clinical benefits of the novel, less toxic PARP inhibitors, and highlight the importance of the inclusion of systems-level toxicities in mathematical cancer therapy models.

## Discussion

PARP inhibitors have yielded significant clinical benefits in the first-line maintenance treatment of high-grade serous ovarian cancer. However, the widespread clinical adoption of PARPi maintenance has given rise to several unanswered questions regarding this therapy and patient management. These questions encompass the optimal duration of olaparib administration, the appropriate dosage scale of the medication, and whether the measures taken to manage toxicity impact the clinical benefits derived from the treatment. Additionally, the dynamics of tumors during PARP inhibitor maintenance remain unknown, posing challenges post the development of therapy resistance. To address these uncertainties, we have developed a computational platform comprising both a mechanistic model and statistical analyses. By employing a virtual clinical trials approach, we demonstrate that heterogeneous tumor dynamics and toxicity management significantly influence clinical outcomes. Our findings underscore the efficacy of virtual clinical trials in optimizing treatment strategies and provide valuable insights for guiding the design of future clinical trials.

Even though olaparib has shown efficacy in patients with *BRCA1/2* mutated/HRD-positive tumors, not all patients respond uniformly well. This heterogeneity is apparent even in clinical trials, where close to 30% of the patients relapse during PARPi maintenance due to emerging PARP inhibitor resistance. Numerous resistance mechanisms to PARP inhibitors have been identified (29). Importantly, resistance to platinum-based chemotherapies is a strong predictor of PARPi resistance, suggesting shared underlying mechanisms (30). In general, a secondary mutation in *BRCA1/2* restoring the protein function is the best clinically characterized mechanism of platinum agent and PARPi resistance in HGSC (31–35). In addition, several non-genetic mechanisms have been identified, including the downregulation of DNA repair proteins (36–38) and enhanced replication fork protection (39–42). Interestingly, previous work has suggested that PARPi resistance is associated with significant clonal, genomic, and functional heterogeneity (43). In alignment with the clinical observations regarding the inferior 2nd PFS in the clinical setting, we observed inferior responses to subsequent therapy after relapse on PARPi treatment, confirming that our mathematical model adeptly reproduces the cancer evolution biology and clinical behavior of overlapping PARPi and platinum therapy resistance mechanisms. Our study presents cancer cell fitness, chemotherapy response, and selection pressure as predictive markers of the response to olaparib, forming a robust foundation for the development of predictive combination biomarkers. Furthermore, our model emphasizes the role of hematological toxicity parameters as determinants of clinical benefit in first-line maintenance treatment with PARP inhibitors.

Even though olaparib is generally well-tolerated, a substantial number of patients report mild or moderate side effects, including fatigue, nausea, and other gastrointestinal symptoms. Many of these issues can be effectively managed with supportive therapy or dose modifications and do not require pausing the treatment. The most prevalent grade 3–4 adverse effects include anemia and neutropenia (44), frequently necessitating treatment interruption and dose reduction. In the SOLO-1 trial, adverse effects led to treatment interruption in 51.9% of olaparib patients, dose reduction in 28.5%, and treatment discontinuation in 11.5% of patients (45). Our findings show that treatment interruptions or dose reductions did not compromise the maintenance treatment outcomes and that dose reductions, even to low doses, should be prioritized over treatment discontinuation. This aligns with previous studies concerning olaparib interruptions in the recurrent setting (46). Our data further support that reductions and interruptions do not compromise the clinical outcomes, providing valuable insights into clinical decision-making and patient counseling concerning toxicity management. Importantly, we show using our computational platform that the use of PARP inhibitors with reduced toxicity results in improved progression-free survival, thereby supporting the development of next-generation molecules with reduced toxicity profiles for maintenance treatments.

Our novel computational platform considers not only the response to drugs but also drug resistance and toxicity. The platform is generic and freely available, and thus, with minimal changes, it could be applied to test other anticancer drugs, facilitating the design of new clinical trials. However, the platform is not without disadvantages. The branching process model does not include the interaction between cancer cells and all other cells in the tumor. In fact, all cancer cells have the same dynamics, regardless of the number of cells present in the tumor. However, it is impossible to properly measure the interactions between cancer cells at the patient scale. Secondly, the developed simulator is based on the assumption that all cancer cells are synchronized (i.e., all of them die and divide at the same time). This allows for model simulation in a reasonable time because the branching process model is a stochastic model that is difficult to simulate numerically. The development of a simulator with an asynchronous mode is left as future work. Lastly, a branching process model is a well-mixed model; thus, we do not simulate the structure of the tumor due to computational limitations.

Altogether, our findings underscore the efficacy of virtual clinical trials that include modeling of resistance mechanisms, tumor dynamics, and patient toxicities in optimizing treatment strategies and provide valuable insights to guide clinical treatment and future clinical trial design.

## Data Availability

All data produced in the present work are contained in the manuscript

## Acknowledgments

Calculations were performed on the Ziemowit computer cluster in the Laboratory of Bioinformatics and Computational Biology, created in the EU Innovative Economy Programme POIG.02.01.00-00-166/08 and expanded in the POIG.02.03.01-00-040/13 project. This work was carried out in part by the Silesian University of Technology internal research funding. The work was supported by grants from the Sigrid Jusélius Foundation (A.F.), Cancer Society of Finland (A.F.), Academy of Finland (grant number 339805, 350396 to A.F.), The Finnish Medical Foundation (A.F.), University of Helsinki (A.F.), and Helsinki University Hospital state-level funding (A.F.). This study was co-funded by the European Union (ERC, SPACE, 101076096). Views and opinions expressed are, however, those of the author(s) only and do not necessarily reflect those of the European Union or the European Research Council. Neither the European Union nor the granting authority can be held responsible for them.

