## Supplementary material for "Mathematical modeling suggests improved clinical outcomes of second-generation PARP inhibitors with reduced toxicity"

January 29, 2024

### Contents


**Text S1.** SOLO-1 clinical trial data 2

**Text S2.** SOLO-2 clinical trial data 4

**Text S3.** Hematological toxicity data 5

**Text S4.** Toxicity management data 6

**Text S5.** Mathematical model 7

**Text S6.** Computational framework integrating patient data with mathematical model 14

**Text S7.** The model parameters and their values justification 15

**Text S8.** Standard-of-care simulation 20

**Text S9.** Computer simulator 23

**Text S10.** Features extracted from virtual SOLO-1 clinical trial simulations 25

### Text S1. SOLO-1 clinical trial data

The SOLO-1 is a phase III clinical trial that tests the application of olaparib as a maintenance treatment in advanced ovarian cancer with *BRCA1/2* mutation. It is a successful clinical trial as based on it, FDA (in 2014) and EMA (also in 2014) approved the olaparib as monotherapy maintenance after the primary treatment with platinum-based chemotherapy.

Figure 1 shows the treatment protocol of patients enrolled to the SOLO-1 clinical trial and eligibility criteria to the trial. Below, we describe in more details this trial and what type of data were utilized from it in the virtual clinical trial simulations we performed.

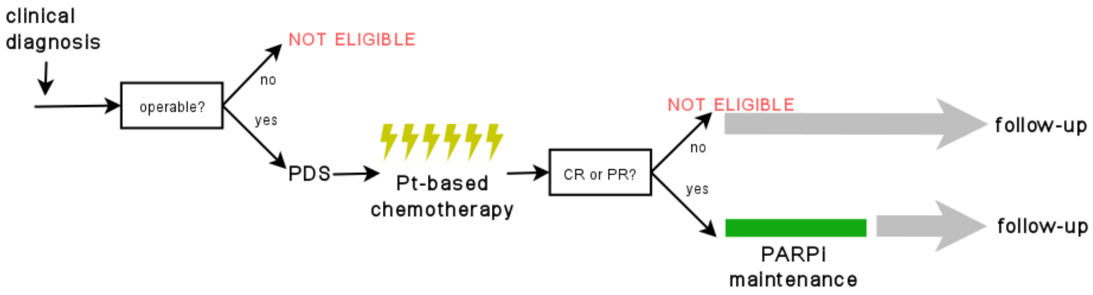


Figure 1: SOLO-1 clinical trial eligibility and protocol schematic.

#### Eligible patients

Those patients with newly diagnosed, advanced, *BRCA1/2*-mutated high-grade ovarian, endometroid, fallopian tube, or peritoneal cancer were eligible to the SOLO-1 clinical trial. All patients underwent at least three cycles of platinum-based chemotherapy with complete response (CR) or partial response (PR) and cytoreductive surgery before administration of olaparib.

In our model, we also focus on high-grade ovarian cancer treated with upfront cytoreductive surgery and platinum-based chemotherapy (carboplatin + paclitaxel). We only consider advanced (with stage III and IV) patients what in the model is measured with tumor burden at the time of diagnosis (defined with parameter $M_{diagnosis}$).

#### Trial design

The SOLO-1 clinical trial is a randomized controlled trial (RCT). Each patient after primary treatment including cytoreductive surgery and platinum-based chemotherapy is assigned to the treatment and placebo group randomly with a 1:1 ratio.

Next, treatment group patients received 150 mg of olaparib twice daily and rest of them received placebo. The maintenance treatment was continued until tumor progression or severe toxcity is observed. In the case of no recurrence after two years from the start of treatment with placebo/olaparib, the treatment is stopped. Thus, each patient is treated maximum two years.

In our, model, we performed the trial in such a way that it resembles the real SOLO-1 clinical trial. Thus, we also randomized patients into placebo/olaparib arms. We also measured if cancer reoccurred or not, by measuring tumor burden. If no cancer is observed after two years, we stopped the simulation and treated the patient as cured one. The difference between real and virtual SOLO-1 trial is a measurement of recurrence. In real SOLO-1, the recurrence is measure using the level of CA-125 biomarker and in virtual SOLO-1 trial through tumor burden measured radiologically.

More importantly, the Kaplan-Meier estimator of first progression-free survival, which measures a patient's response to the treatment used, was obtained from the SOLO-1 clinical trial.

#### Outcome

The primary outcome measured in the SOLO-1 trial is progression-free survival (PFS) which is measured as the time interval (in months) between trial randomization and cancer recurrence.

In the SOLO-1 clinical trial, the time to recurrence is measured using radiological scans and CA-125 blood biomarker. In the case of the mathematical model, we measure tumor burden to estimate when patients relapse. We assume that the tumor burden is a reliable metric of cancer recurrence.

We applied the Kaplan-Meier statistical model to estimate the fraction of patients in remission as a function of time. In such a way, for the whole population of virtual and real SOLO-1 clinical trial, we know the distribution of PFS. From the simulation, we extracted the PFS for each virtual patient as described above. PFS from a real SOLO-1 clinical trial is not available. However, there is an available Kaplan-Meier plot for placebo/olaparib treatment arms [10]. Thus, we extracted the Kaplan-Meier curves using DataThief software [16].

### Text S2. SOLO-2 clinical trial data

SOLO-2 trial evaluates olaparib in relapsed ovarian cancer that was previously treated with chemotherapy. This trial tests olaparib versus placebo in ovarian cancer patients. The goal is to evaluate the chemotherapy following disease progression on patient treated with olaparib maintenance treatment. It is double-blind, randomised, placebo-controlled, and phase 3 trial.

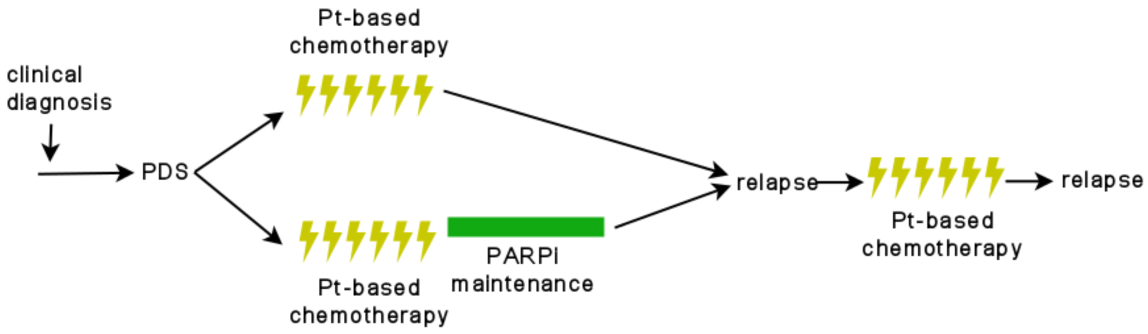


Figure 2: SOLO-1 clinical trial eligibility and protocol schematic.

#### Eligibility criteria

To SOLO-2 the patients with following criteria could be enrolled:

1. with high-grade serous ovarian cancer or high-grade endometrioid cancer, including primary peritoneal or fallopian tube cancer,
2. platinum-sensitive,
3. ECOG score 0 or 1,
4. relapsed after first-line treatment.

In summary, those patients who received chemotherapy and olaparib or chemotherapy alone and are relapsed are eligible. It is the same scenario as in our virtual clinical trial.

#### Trial design

This trial is designed as follows. Only those patients that relapse after first-line treatment are eligible leading to a smaller cohort of patients both in real data and in simulations.

All relapsed patients received 6 cycles of Pt-based chemotherapy. No additional treatment is administered. After chemotherapy, patients are follow-up until the second relapse.

### Text S3. Hematological toxicity data

To fit the toxicity parameters of the developed model, we applied temporal data from Yamamoto 2002 et al. [18]. Those data are from phase I/II clinical trial where the goal was to investigate toxicity and efficacy of carboplatin-paclitaxel chemotherapy doublet.

The toxicity data includes, among others, the level of white blood cells (WBC) in the patient's blood during the first-line chemotherapy cycles. Estimated values, together with the time when WBC's level is the lowest after each cycle of chemotherapy (nadir), are presented in Table S1.

The normal level of WBC in an adult woman's body is in range of $4.5 - 11.0 \cdot 10^9/L$. After the platinum-based chemotherapy cycle, when the level of Leukocytes drops to $2.0 \cdot 10^9/L$ it is considered as the toxicity occurence. The lowest value of Leukocytes is observed after about 10 days.

Table S1: Hematological toxicity measured by Yamamoto 2002 et al. [18]. Each column shows the results from one chemotherapy cycle. The lowest value of WBC and nadir, adapted from [18], are presented.

| | 1 | 2 | 3 | 4 | 5 | 6 |
|---|---|---|---|---|---|---|
| The lowest value [$10^9/l$] | 2.3 | 2.1 | 2.1 | 2.0 | 1.9 | 2.0 |
| Nadir [days] | 10 | 10 | 10 | 10 | 9.5 | 10 |

We applied the data to calibrate the counts of WBC during chemotherapy. The data about WBC concentration were scaled to WBC level in adult human body. This data were applied to estimate the level of toxicity of platinum-based chemotherapy (parameter $\gamma_{pt-based\ chemotherapy}$).

### Text S4. Toxicity management data

From SOLO-1 clinical trial, we extracted information about adverse events. We focused on grade 3-4 adverse events as those require proper management. In the clinical trial, three methods of side effects management were applied:

1. discontinuation - treatment with olaparib is stopped completely,
2. dose reduction - dose of olaparib is reduced,
3. dose interruption - treatment with olaparib is stopped for a given number of days.

Table S2: Percentage of patient with one of three methods of toxicity management.

| **Type of management** | **Percentage** |
|---|---|
| Led to discontinuation of intervention | 12% |
| Led to dose reduction | 28% |
| Led to dose interruption | 52% |
| Total with grade 3-4 adverse events | 39% |

Table S2 shows the data about percentage of patients that required adverse events management. Dose interruption was the most common method for adverse events management. It is important to mention that some patients underwent two or all methods. However, it is impossible to discriminate fraction of those patients from oryginal data provided.

Those data were applied for calibration of virtual patients cohort. As the real patient cohort includes patients that required toxicity management, it should be reproduces in simulation of SOLO-1 clinical trial.

### Text S5. Mathematical model

#### Model structure

The developed mathematical model is a mechanistic model describing tumor growth with and without treatment interventions according to tumor evolution notions (see Figure **Text S5.**). Also, we included in the model the drug resistance as well as toxicity of considered drugs. The model is in form of a discrete-time multiple-type branching process model with two types of cells: cancer cells and white blood cells (WBC). We included cancer cells as a heterogeneous mass of cells, whereas WBC cells as a homogeneous one.

We consider that each cancer cell is characterized by a level of resistance to platinum-based chemotherapy and PARP inhibitor (PARPi) - olaparib. Thus, each cancer cell is defined with two numbers, where the first one defines the number of resistance mechanisms accumulated to pt-based chemotherapy and the second one the number of resistance mechanisms accumulated to PARPi. This way, we model tumor as a heterogeneous mass of cancer cells where number of drug resistance mechanisms accumulated by cancer cell can in theory go to infinity.

WBC population, however, is homogeneous. Thus, WBC population is modeled as one-type branching process. In contrast to cancer cells where we assume they grow exponentially, WBC is assumed to grow logistically with carrying capacity $K$. We also assumed that cancer cells do not interact with WBC. Instead, the population of WBC is a biomarker for the toxicity of anti-cancer treatment.

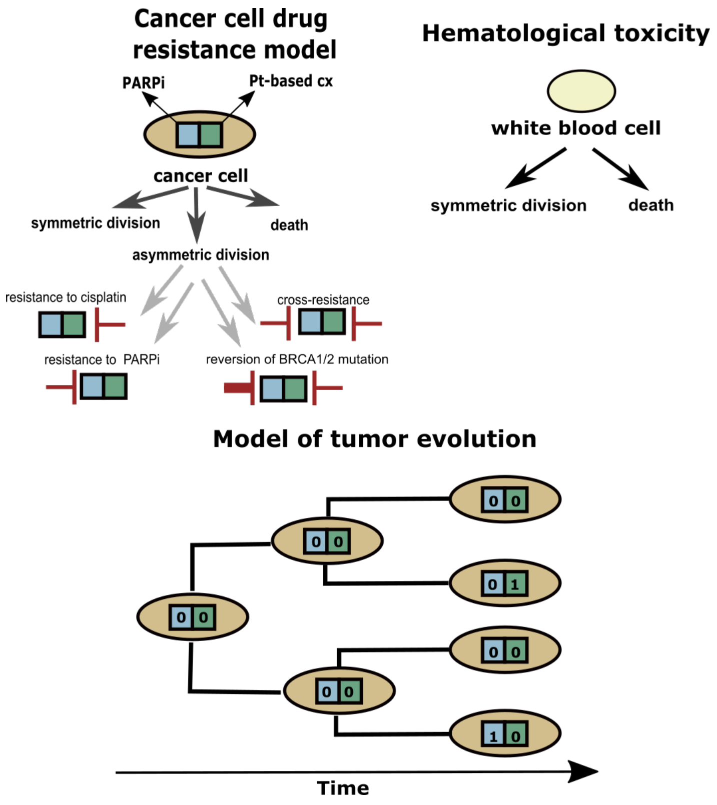


Figure 3: The structure of the model. Cancer cells are described using multiple-type branching process where each subclone is characterized with different level of drug resistance to platinum-based chemotherapy and olaparib. White blood cells are describe with a single-type branching process. In addition to developing additional drug resistance to olaparib/platinum-based chemotherapy, so-called reversion mutation and cross-resistance may also happend.

#### Evolution of drug resistance to anti-tumor agents

It is known that the acquisition of resistance to both types of treatments which are considered herein is an evolutionary process. There are two well-described models of drug resistance evolution - stochastic and hierarchical one. The first model hypothesizes that the resistance to treatment in cancer is a result of the acquisition of (epi)genetic aberration by cancer cells. These aberrations lead to drug resistance. The second model assumes that there are a small fraction of cancer cells which are called cancer stem cells (CSC) that have self-renewal potential. Next, those cells differentiate leading to the appearance of differentiated cells that are the most frequent cancer cells in the tumor. It is hypothesized that CSCs are cells that acquired already the resistance to anticancer treatment.

Herein, we chose a stochastic model of drug resistance evolution. The rationale for this choice is the fact that ovarian cancer patients acquire resistance stochastically [9]. Also, there is strong evidence that resistance to pt-based chemotherapy and PARPi appear before treatment [4]. Thus, here we focus on pre-existing drug resistance.

Ovarian cancer is characterizes by frequent copy number variation (CNV) and rare point mutations. Thus, instead of the number of mutations connected with drug resistance, we talk about mechanisms of resistance which are results of the acquisition of both CNVs and point mutations. Thus, a cancer cell in the mathematical model is described with a number of resistance mechanisms to pt-based chemotherapy and olaparib (PARPi). For example, a cancer cell depicted as $[2, 1]$ has two resistance mechanisms to pt-based chemotherapy and one to olaparib accumulated.

In the absence of treatment intervention, resistant cells (all cells except the wild-type $[0, 0]$) behave as wild-type cells. Thus, we consider the neutral evolution model as the growth rate of both types of cells is identical. However, during the treatment, the growth rate is dependent on the number of drug resistance accumulated as shown in Figure 4.

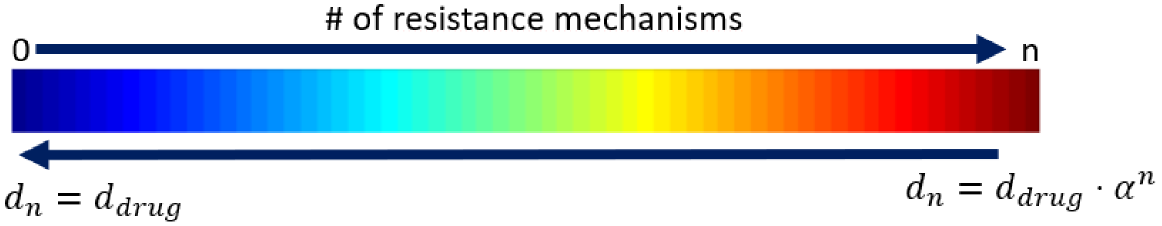


Figure 4: The model of drug resistance, which is a discrete trait in our model. Number of drug resistance mechanisms accumulated is inversely-correlated with drug-induced death rate ($d_{drug}$) and $0 < \alpha < 1$.

As shown in Figure 4, when the number of drug resistance mechanisms increases, the drug-induced death rate ($d_{drug}$) decreases exponentially with rate $\alpha$, where $\alpha$ is always below one. In the case when two drug resistance mechanisms to one drug are accumulated in the cell, the $d_{drug}$ equals:

$$d_n = d_{drug} \cdot \alpha \cdot \alpha = d_{drug} \cdot \alpha^2 \tag{S1}$$

leading to multiplicative model of resistance.

Without loss of generality, we can assume that each mechanism of drug resistance is equal and leads to the same reduction of a drug-induced death rate. This also reduces the number of model parameters. In the case of two drugs, the drug-induced death rate equals:

$$d_{drug} = d_{drug_1} \cdot \alpha_1^{n_1} + d_{drug_2} \cdot \alpha_2^{n_2} \tag{S2}$$

#### Anti-tumor treatment

We included in the model both the efficacy and pharmacokinetics of the anticancer agents. This allows us to model drug concentration over time after drug administration. This, in turn, helps to optimize the treatment more accurately.

Below, we describe three types of treatment modalities included in the model: cytoreductive surgery, platinum-based chemotherapy, and targeted treatment using the PARP inhibitor called olaparib. Each treatment modality has a different goal. Debulking surgery is performed to remove as many cancer cells as possible locally at one-time point. Chemotherapy is performed in an adjuvant setting to kill remaining cancer cells. Lastly, olaparib is administered as maintenance therapy to prolong progression-free survival by preventing cancer regrowth.

##### Debulking surgery

Cytoreductive surgery is a type of surgery performed to resect the whole tumor. This type of surgery in ovarian cancer takes about 6 hours. As we simulate the model for a time-frame of months, we can assume that cytoreductive surgery is the removal of a fraction of cancer cells at one time-point. Next, we don't know which type of cancer cell will be removed at the time of surgery. Thus, we can also assume that each cell, despite its type, is removed with the same probability leading to intact cancer cells composition. In the model, we included cytoreductive surgery by removal of $\beta$ cancer cells from each subtype of cancer cells at a one-time point.

##### Pt-based chemotherapy

Platinum-based chemotherapy includes carboplatin + taxane (paclitaxel) drug combination and is administered as intravenous (IV) bolus injection. The injection of diluted carboplatin combined with taxane takes about 6 hours. As the main agent in the chemotherapy mixture is carboplatin, we model only concentration and the effect on this agent. Because of the short time, it takes to infuse the chemotherapy in comparison with the time we are simulating, we assume that the carboplatin reaches maximal concentration in the tumor at the one-time point when the drug is administrated. After injection, the drug concentration decays exponentially according to the following algebraic equation describing the pharmacokinetics of carboplatin:

$$C_{pt-based\ chemotherapy}(t+\Delta t) = C_{pt-based\ chemotherapy}(t) \cdot e^{-k_{pt-based\ chemotherapy} \cdot t}. \tag{S3}$$

The effect of chemotherapy is modeled as follow. In the model, we included an additional death rate called chemotherapy-induced death rate which is different for each subclone as mentioned in the 'Evolution of resistance to anti-tumor agents' subsection. The chemotherapy-induced death rate equals:

$$d_{pt-based\ chemotherapy}(t+\Delta t) = b \cdot C_{pt-based\ chemotherapy}(t), \tag{S4}$$

where $b$ is the birth rate. Thus, the effect of pt-based chemotherapy is included according to the Norton-Simon hypothesis which states that the rate of cancer cell death in response to treatment is directly proportional to the tumor growth rate at the time of treatment. As a result, the same fraction of cancer cells is killed no matter what is the initial amount of cancer cells.

##### Olaparib

Olaparib, in contrast to platinum-based chemotherapy, is administered orally in form of tablets. Usually, the olaparib is administered with a dose of 150 mg twice a day. This gives us a daily dose of 300 mg. The time to maximum drug concentration ($t_{max}$) is one hour [15]. Thus, similarly to modeling pt-based chemotherapy concentration, we assume that the maximal concentration ($C_{max_{PARPi}}$) is immediately after drug administration. Next, according to the one-compartmental pharmacokinetic model, the olaparib concentration decreases exponentially. Thus, olaparib concentration is given with the following algebraic equation:

$$C_{PARPi}(t + \Delta t) = C_{PARPi}(t) \cdot e^{-k_{PARPi} \cdot t}. \tag{S5}$$

The effect of targeted therapy is modeled as followed. In the model, we included an additional death rate called drug-induced death rate which is different for each subclone as mentioned in the 'Evolution of resistance to anti-tumor agents' subsection. The drug-induced death rate equals:

$$d_{PARPi}(t + \Delta t) = b \cdot C_{PARPi}(t), \tag{S6}$$

where $b$ is the birth rate. Thus, the effect of olaparib is included according to the Norton-Simon hypothesis similarly to platinum-based chemotherapy.

#### Hematological toxicity

The last element of the model is inclusion of toxicity. As myelosupression which lead to leukophenia is dose-limiting type of drug toxicity, we model the dynamics of white blood cells as a biomarker of treatment toxicity. In absence of treatment intervention, the amount of blood cells oscillate around carrying capacity value $K$. Here, the amount of WBC is govern with the following algebraic equation:

$$WBC(t + \Delta t) = WBC(t) \cdot (b(t) - d(t)) \tag{S7}$$

where $b(t)$ is birth rate and is equals:

$$b(t) = \frac{1}{2}\left(1 + s_{WBC}\left(1 - \frac{WBC(t)}{K_{WBC}}\right)\right) \tag{S8}$$

and $d(t)$ is death rate equals:

$$d(t) = 1 - b(t). \tag{S9}$$

In the case when $WBC(t) = K_{WBC}$, the WBC has the same probability to divide and die as $b(t) = d(t) = \frac{1}{2}$. When $WBC(t) << K_{WBC}$, $b$ has the highest value what allow exponential growth of WBC. Together with the increase in the number of WBC in the system, the birth rate decrease, and the death rate increase.

Now, adding the treatment intervention, we include drug-induced WBC death. Here, number of WBC is govern with the following equation:

$$WBC(t + \Delta t) = WBC(t) \cdot (b(t) - d(t) - d_{pt-based\ chemotherapy_{WBC}}(t) - d_{PARPi_{WBC}}(t)) \tag{S10}$$

where: $d_{pt-based\ chemotherapy_{WBC}} = \gamma_{pt-based\ chemotherapy} \cdot WBC(t)$, and $d_{PARPi_{WBC}} = \gamma_{PARPi} \cdot WBC(t)$. Thus, the toxicity in the model is described with two parameters: $\gamma_{pt-based\ chemotherapy}$ and $\gamma_{PARPi}$, which are called drug-induced death rate of WBC. As a result of drug toxicity, number of WBC decrease.

The inclusion of drug toxicity in the mathematical model allows us to better control the dosing of chemotherapy and targeted treatment. Firstly, it will exclude unrealistic scheduling like maximum tolerated dose scheduling with very small intervals between cycles. Secondly, it is a known problem that pt-based chemotherapy and PARPi cannot be administered at the same time due to drug toxicity. Thus, the inclusion of toxicity removes from all possible schedules those, which assume administration of pt-based chemotherapy together with PARPi. Lastly, it allows optimizing the drug schedule in such a way that drug concentrations and time of its administration is balanced between toxicity and drug effectiveness.

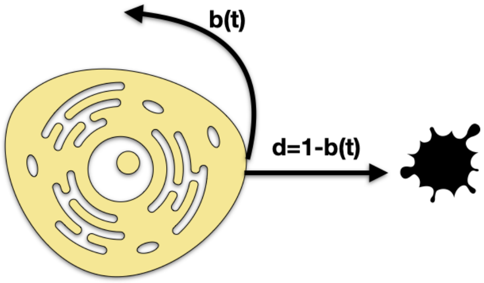


Figure 5: Inclusion of hematological toxicity into the mathematical model.

#### Key assumptions of the model

In the Table below, we listed all important assumptions of the mathematical model.

Table S3: Model assumptions. Assumptions marked as 'simplification' are created to simplify the model.

| **assumption** | **reference** |
|---|---|
| **Assumptions related to growth of cancer cells** | |
| All cancer cells are synchronized, and thus are dividing at the same time | simplification |
| A cancer cell is defined as a sequence of two numbers where each place is a different drug and the value means how resistant to a given drug the cancer cell is | simplification |
| An ovarian tumor grows exponentially | biological knowledge |
| All cancer cells are growing with the same net growth rate in absence of treatment what is in line with the neutral cancer evolution model | [17] |
| At each division, a cancer cell can acquire one resistance mechanism to a given drugs | [8] |
| The growth of each cancer cell type is independent of other cancer cells, meaning that we do not consider the interaction between the subclones | simplification |
| **Assumptions related to cancer treatment and its toxicity** | |
| Surgery is performed as removal of a tumor at a one-time point | clinical practice |
| Surgery leads to the removal of each of the cancer cell with the same probability | clinical practice |
| Pharmacokinetics of pt-based chemotherapy and PARPi is included as one-compartmental drug decay model | simplification |
| Chemotherapy is administered with maximum tolerated dose (MTD) | clinical practice |
| Drug administration leads to non-zero drug-induced death rate | simplification |
| Treatment leads to toxicity measured as as a drop of the WBC level in the blood | clinical practice |
| Surgery does not lead to a decrease in WBC | simplification |
| pt-based chemotherapy is administered as an IV bolus injection and olaparib orally | clinical practice |
| **Assumptions related to cancer treatment resistance** | |
| Each subclone in the model vary with the level of drug resistance accumulated | biological knowledge |
| Drug resistance leads to a decreased drug-induced death rate of a cancer cell | simplification |
| The number of resistance mechanisms can theoretically go to infinity | simplification |
| Acquisition of resistance is irreversible in case of pt-based chemotherapy | simplification |
| Acquisition of resistance to olaparib leads also to resistance to pt-based chemotherapy | biological knowledge |

| | |
|---|---|
| During cell division, cancer can acquire one resistance mechanism to only one drug | simplification |
| **Miscellaneous assumptions** | |
| Cancer initiation starts from a single sensitive cell | simplification |
| Cancer is diagnosed when the number of cancer cells crosses some threshold | clinical practice |
| There is no delay between cancer diagnosis and the start of treatment | simplification |
| Cancer relapse is detected when tumor burden is above 1 $cm^3$ which is the smaller detectable tumor using CA-125 biomarker | clinical practice |

### Text S6. Computational framework integrating patient data with mathematical model

The mathematical model developed herein is integrated with the data from two clinical trials that are described in **Text S1** and **Text S2**. The data from this trial concerning pharmacokinetics, drug toxicity, drug resistance, and response rate were applied to parametrize the model. Next, we applied the outcome of the model (progression-free survival times) to create survival curve using Kaplan-Meier estimator. The curve is then used to verify the accuracy of the mathematical model by comparing with Kaplan-Meier from SOLO-1 clinical trial.

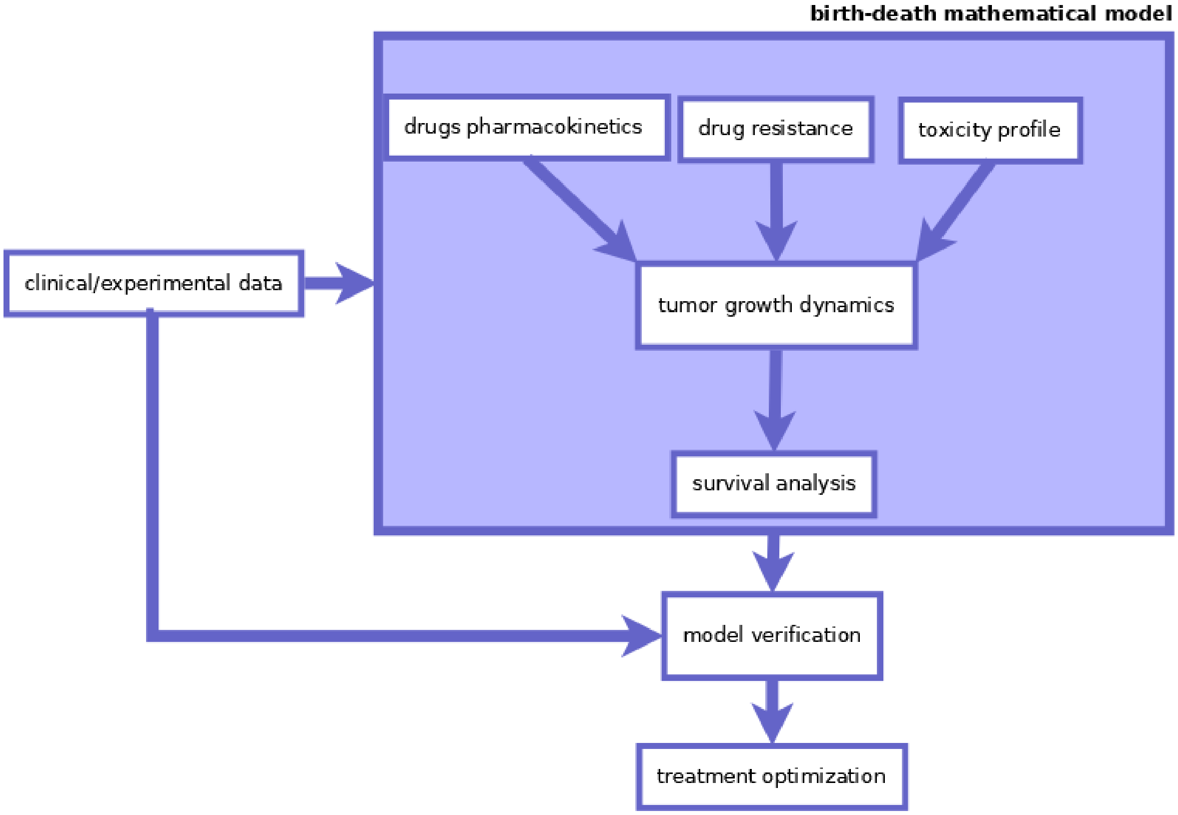


Figure 6: The computational pipeline integrating mathematical model and three types of data: PK, drug resistance, toxicity profile.

In Figure 6, we present the pipeline of integration data with the mathematical model. The data from SOLO-1 clinical trial and phase I/II clinical trial of platinum-based chemotherapy [18] are collected. Next, the data are processed to extract all necessary mathematical model parameters. The parameters could be divided into three categories: drug pharmacokinetics, drug resistance, and drug toxicity. Those parameters are input to the birth-death branching process model which is described in detail in **Text S3**. In the next step, the mathematical model is simulated to create a virtual patient cohort which in turn could be applied to create a Kaplan-Meier curve. The model is then verified using the Kaplan-Meier plot from clinical trial patients. The process from parameter extraction to model verification is repeated until convergence. When the model is well-parametrized, we could apply it to optimize the treatment.

The computational framework is developed in MATLAB 2021 environment and available at Github.

### Text S7. The model parameters and their values justification

The Table S4 shows all the model parameters. Some of them were fitted using data from SOLO-1 clinical trial, some using toxicity estimation from [18] and the rest were fitted to clinical knowledge and Kaplan-Meier survival curve.

Figure 7 shows steps-by-step how the developed mathematical model was calibrated. In short, the model parameters related to cancer cell growth and drug resistance accumulation are fitted in the first step. Next, the fitted parameters are set and the parameters related to the pharmacokinetics of the drugs are estimated. Next, drug response parameters are fitted. Using drug toxicity information, we fit the parameters related to pt-based chemotherapy and olaparib toxicity. In the last step, using knowledge from clinical practice, we fitted the rest parameters such as the time between two consecutive olaparib administration.

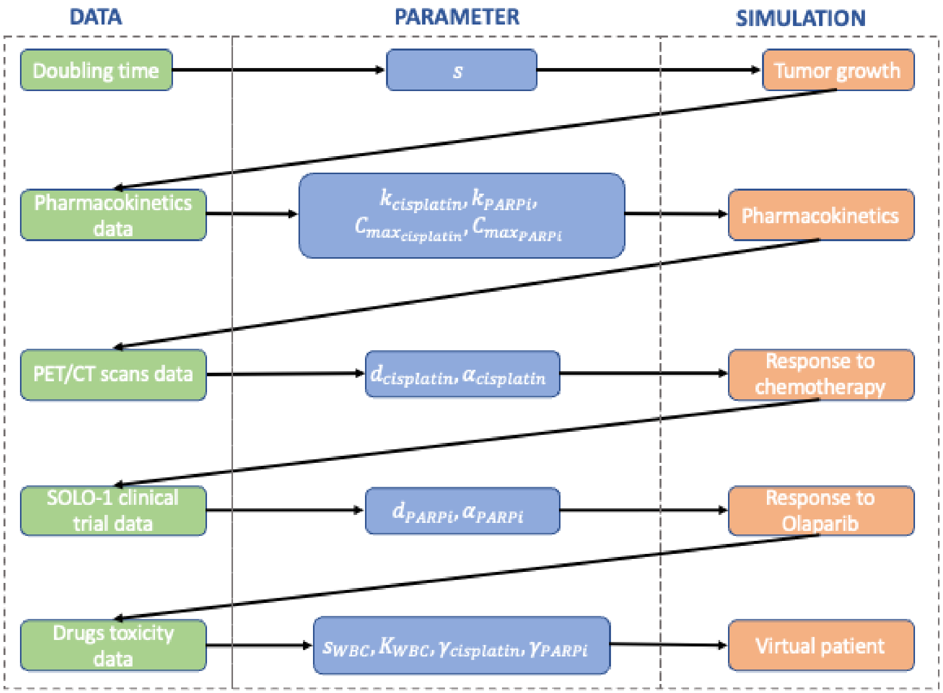


Figure 7: The computational pipeline integrating mathematical model and three types of data: PK, drug resistance, toxicity profile.

The list of mathematical model parameters together with their values is presented in Table S4. There are in total 20 parameters divided into 5 classes.

Table S4: List of model parameters.

| Symbol | Value | Description | Reference |
|---|---|---|---|
| **Cancer and Lymphocytes growth parameters** | | | |
| $s$ | $0.0012 - 0.0194$ | Selection advantage of cancer cells | [12] |
| $s_{WBC}$ | 0.025 | Selection advantage of Lymphocytes | calibrated |
| $K$ | $2.25 - 5.25 \cdot 10^{10}$ | Carrying capacity of Lymphocytes | clinical knowledge |
| **Drug resistance parameters** | | | |
| $u_{pt-based\ chemotherapy}$ | $10^{-9}$ | rate of accumulation of pt-based chemotherapy resistance | [5] |
| $u_{olaparib}$ | $10^{-9}$ | rate of accumulation of olaparib resistance | [7] |
| $u_{cross}$ | $10^{-12}$ | rate of accumulation of cross-resistance in the absence of olaparib | calibrated |
| $u_{crossO}$ | $3 \cdot 10^{-7}$ | rate of accumulation of cross-resistance in the presence of olaparib | calibrated |
| $u_{reversion}$ | $10^{-12}$ | rate of accumulation of cross-resistance in the absence of olaparib | calibrated |
| $u_{reversionO}$ | $2.5 \cdot 10^{-7}$ | rate of accumulation of cross-resistance in the presence of olaparib | calibrated |
| $\alpha_{pt-based\ chemotherapy}$ | 0.5 | decay in pt-based chemotherapy-induced death rate | [6] |
| $\alpha_{olaparib}$ | 0.5 | decay in olaparib-induced death rate | [6] |
| **Pharmacokinetics parameters** | | | |
| $k_{pt-based\ chemotherapy}$ | 0.005 | decay of pt-based chemotherapy | calibrated |
| $k_{olaparib}$ | 0.6850 | decay of olaparib | calibrated |
| $Cmax_{pt-based\ chemotherapy}$ | 459.7 | maximum pt-based chemotherapy concentration after administration | clinical knowledge |
| $Cmax_{olaparib}$ | 300 | maximum olaparib concentration after administration | clinical knowledge |
| **Drug efficacy parameters** | | | |
| $d_{pt-based\ chemotherapy}$ | 0.027 | pt-based chemotherapy-induced death rate of sensitive cells | calibrated |
| $d_{olaparib}$ | 0.004 | olaparib-induced death rate of sensitive cells | calibrated |
| **Drug toxicity parameters** | | | |
| $\gamma_{pt-based\ chemotherapy}$ | $0.6 \cdot 10^{-14}$ | pt-based chemotherapy toxicity rate | calibrated |
| $\gamma_{olaparib}$ | $0.261 \cdot 10^{-14}$ | olaparib toxicity rate | calibrated |

| Miscellaneous parameters | | | |
|---|---|---|---|
| $M_{diagnosis}$ | log-normal distribution with $\mu = 26.593$ and $\sigma = 0.471$ | Tumor burden at diagnosis | [6] |
| $M_{relapse}$ | $10^{10}$ | Tumor burden at relapse | [6] |
| $time_{olaparib}$ | 0.5 [days] | Time interval in hours between two consecutive olaparib doses | clinical knowledge |
| $\beta$ | 0.999 | fraction of cells removed by surgery | [6] |

#### Cancer cells growth dynamics

We assume exponential cancer cell growth and thus we have one parameter to fit - growth rate. As seen in Equation S14, the birth rate is computed using information about selection advantage which is a percentage stating how much bigger is the growth rate of cancer cells in comparison with healthy cells.

The selection advantage is computed according to well known rule of 70, which is an approximation of the percentage from natural logaritm of 2 ($ln(2) = 0.693$). Rule of 70 states that growth rate equals:

$$growth\ rate = \frac{70}{DT} \tag{S11}$$

where $DT$ is doubling time. Next the value obtain with the above equation was divided by $t_{step} \cdot 100$, giving us the following equation for relation between doubling time and selection advantage:

$$s = \frac{70}{DT \cdot n_{step} \cdot 100} = \frac{7}{DT \cdot 20} \tag{S12}$$

where $n_{step} = 2$ is amount of steps within a unit of time (days), as we update the system every 12 hours.

For ovarian cancer, doubling time is estimated to be between 18 and 300 days. Thus, the value of selection advantage is between 0.0012 and 0.0194. In the model simulation, we sample selection advantage from a uniform distribution with ranges defined with the minimum and maximum value of selection advantage.

#### Drug resistance parameters

Drug resistance is described with parameters $u_{pt-based\ chemotherapy}$, $u_{olaparib}$, $u_{reverse}$, and $u_{cross}$ which are describing the rate of accumulation of (epi)genetics aberrations. All the parameters are probabilities of accumulation of one (two) drug(s) resistance mechanism per one cell cycle.

In the case of parameter $u_{pt-based\ chemotherapy}$ and $u_{olaparib}$, we set its value to $10^{-8}\frac{1}{cell\ cycle}$. The values were set through fitting to Kaplan-Meier survival curve for PFS. As we have the first and the second PFS as well as two clinical trial arms, we have in total four Kaplan-Meier survival curves to which those parameters were fitted.

Parameter $u_{cross}$ and $u_{reversion}$, however, is set to $10^{-12}\frac{1}{cell\ cycle}$ in case olaparib is not administered. The value of this parameter is fitted also to Kaplan-Meier survival curves too.

In reality, transition rates are not constant and change over time due to, for example, treatment intervention. Thus, we incorporate this phenomenon into our model by changing value of $u_{cross}$ to $3 \cdot 10^{-7}$ and $u_{reversion}$ to $2.5 \cdot 10^{-7}$ during PARPi maintenance.

#### Tumor burden at the diagnosis

We assume that a tumor is diagnosed when the number of cancer cells crosses some threshold value ($M_{diagnosis}$). Thus, diagnosis is included in the model assuming that diagnosis is made through radiological assessment. It was previously shown that tumor burden at the diagnosis ($M_{diagnosis}$) follows log-normal probability distribution. Thus, here, for each virtual patient, the value of $M_{diagnosis}$ is drawn from a log-normal distribution with $\mu = 26.593$ and $\sigma = 0.471$. These parameters are estimated using a cohort of 28 patient with high-grade serous ovarian cancer who underwent PET/CT scans at the time of diagnosis [6]. Without loss of generality, we assume that the time between diagnosis and performance of PET/CT scan is equal to zero.

#### Pharmacokinetics parameters

There are two types of pharmacokinetics parameters in the mathematical model: maximal drug concentration ($C_{max}$) and decay rate of a drug in the cancer cells ($k$). As we include in the model two drug, we have four pharmacokinetics parameters: $C_{max_{pt-based\ chemotherapy}}$,$k_{pt-based\ chemotherapy}$, $C_{max_{olaparib}}$,$k_{olaparib}$, .

Without loss of generality, we can assume that $C_{max}$ in our mathematical model equals the drug dose administered to the ovarian cancer patients. Thus, for olaparib, $C_{max} = 300$ $[mg]$ which is a dose administered in SOLO-1 clinical trial. For pt-based chemotherapy, $C_{max}$ equals to 5 AUC dose which could be transferred to dose in mg using Calvert equation [3]:

$$dose[mg] = AUC \cdot (GFR + 25) \tag{S13}$$

where $GFR$ is the glomerular filtration rate. Equation S13 takes information about kidney function. To compute $GFR$, we need information about: patient weight, level of creatinine, and patient age. Patient age we have chosen as the median age at which ovarian cancer is diagnosed. Next, for this age, we have found the mean weight for females. Lastly, the median value of creatinine of patients with functioning kidneys is estimated. Table S5 shows the values of each of those parameters necessary for the calculation of pt-based chemotherapy dose. After the substitution of all necessary parameters to the Clavert equation, we obtained a maximal dose of pt-based chemotherapy equal to $C_{max_{pt-basedchemotherapy}} = 459.7[mg]$.

Table S5: Patient parameters for calculation dose of pt-based chemotherapy.

| **parameter** | **value** | *reference* |
|---|---|---|
| weight | 166.5 lb | [1] |
| age | 63 years | [2] |
| creatinine | 90.65 $\frac{\mu mol}{L}$ | [14] |

Next, $k_{pt-based\ chemotherapy}$ and $k_{olaparib}$ were fitted as follows. From the DrugBank database, we extracted the so-called terminal half life of pt-based chemotherapy and olaparib. Knowing the terminal half-life of pt-based chemotherapy (10 days) and olaparib (0.5 days), we can easily fit the $k_{pt-based\ chemotherapy}$ and $k_{olaparib}$ in such a way that after administration of a single dose of a drug, after half-time life, the concentration decrease to half of an initial dose. As a result, we get the $k_{pt-basedchemotherapy} = 0.005[1/day]$ and $k_{olaparib} = 0.6850[1/day]$.

#### Treatment response parameters

There are five treatment parameters in the model: $d_{pt-based\ chemotherapy}$, $d_{PARPi}$, $\alpha_{pt-based\ chemotherapy}$, $\alpha_{olaparib}$ and $\beta$. The first two parameters describe the drug-induced death rate on sensitive cells, the third and fourth parameter describes the decay rate of death rate as a result of drug resistance. The last parameter describes cytoreductive surgery and is defined as a fraction of cancer cells removed by surgery. In the model, $\alpha_{pt-based\ chemotherapy} = \alpha_{olaparib} = 0.5$ as we assume that each additional level of the resistance mechanism of a particular drug reduce a corresponding drug efficacy by half. Next, parameter $\beta$ is taken from [13] where it is estimated that surgeons remove about 99.9% of cancer cells.

We fitted the parameters $d_{pt-based\ chemotherapy}$ and $d_{olaparib}$ as follows. From our previous work, we know that 3 cycles of Pt-based chemotherapy lead to (on average) a 1-log-kill reduction of the tumor [6]. Thus, 6 cycles lead to 2 log-kill reductions of the tumor. As a result, 99% of cancer cells are removed by primary platinum-based chemotherapy. We fitted $d_{pt-based\ chemotherapy}$ in such a way that (on average) chemotherapy leads to the removal of 99 % of the simulated cancer cells. In summary, $d_{pt-based\ chemotherapy}$ was set to 0.027, and $d_{PARPi}$ was set to 0.004 as a result of fitting the parameter to the Kaplan-Meier plot of SOLO-1 clinical trial.

#### Hematological toxicity parameters

Parameters related to hematological toxicity were fitted to data from Yamamoto (2002) et al. [18] and SOLO-1 clinical trial. The parameters $s_{wbc}$ and $\gamma_{pt-based\ chemotherapy}$ were fitted in such a way that the toxicity profile of pt-based chemotherapy fits the observed one. With values of $s_{wbc} = 0.025$ and $\gamma_{pt-based\ chemotherapy} = 0.6 \cdot 10^{-14}$ we obtained the best fit of the model with toxicity profile.

The third parameter describing the toxicity profile is $\gamma_{olaparib}$ describing toxicity to olaparib. 40% of patients have a toxicity profile that required toxicity maintenance. Thus, $\gamma_{PARPi}$ was fitted in such a way that 40% of patients have toxicity to olaparib, so $\gamma_{olaparib} = 0.261 \cdot 10^{-14}$.

#### Miscellaneous parameters

Tumor burden at the diagnosis was fitted to PET/CT scans data retrieved from Kozłowska et al. 2018 [6]. There, metabolic tumor volume (MTV) was estimated as what could be treated as the number of cancer cells at diagnosis. Based on the patient cohort, we fitted the number of cancer cells to various probability distributions and obtained the best fit for log-normal distribution.

Tumor burden at relapse was also taken from Kozłowska et al. 2018. It was justified there that tumor recurrence is observed through CA-125 biomarker level when the amount of cancer cells is above $10^{10}$ which is about 10 $cm^3$.

### Text S8. Standard-of-care simulation

The treatment simulator consists of five phases: pre-treatment, first-line treatment, maintenance treatment, second-line treatment, and treatment-free phase. The first phase starts with cancer initiation and ends with a cancer diagnosis. The second phase contains the primary treatment of ovarian cancer patients. The third phase is maintenance treatment with olaparib until the first relapse.The last two phases of simulation is the second-line chemotherapy as well as resting phase after the second-line treatment if patient will not relapse during treatment phase.

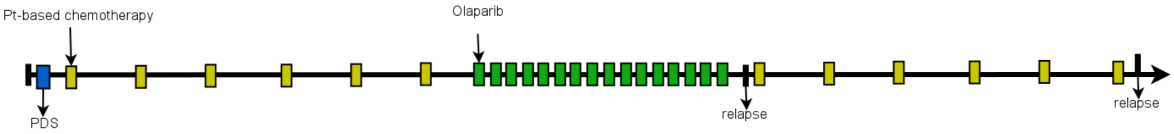


Figure 8: Schematic of standard of care for advanced high-grade serous ovarian cancer.

Figure 8 shows the structure of standard-of-care for advanced ovarian cancer. Figure 9, show the diagram of the simulator of high-grade serous ovarian cancer developed herein and available at Github `add_later_link`.

#### Pre-treatment phase

The simulation starts from the number of Lymphocytes at carrying capacity level and a single fully-sensitive cancer cell. Thus, simulations start from a cancer initiation. In this phase of simulation, the tumor grows without any treatment perturbation until the diagnosis, which is defined in our model as a time when the amount of tumor cells is above some threshold ($M_{diagnosis}$). It means that diagnosis in our simulations is performed radiologically.

Pre-treatment phase is performed to get proper cohort of diagnosed patients that differ in the response to treatment, level of drug resistance, and toxicity profile.

#### Treatment phase

The treatment phase starts at the time of cancer diagnosis. Without loss of generality, we can assume that the patient starts treatment on the same day when is diagnosed. The primary treatment includes both up-front cytoreductive surgery and platinum-based chemotherapy.

Primary debulking surgery (PDS) is administered by the removal of a fraction of cancer cells at a one-time point. After surgery, there is a resting phase which is 30 days long. The break is incorporated to take into account that chemotherapy does not start immediately after surgery but with a delay because it takes time to recover from this aggressive cytoreductive surgery. Next, six cycles of platinum-based chemotherapy are administered with a median time interval of 21 days. The time to the next cycle is defined by the level of Lymphocytes. If the Lymphocyte level is below given threshold, the chemotherapy cycle is postponed for one week.

After the sixth cycle of platinum-based chemotherapy, there is a resting phase during which, the level of WBC comes back to its normal level. The goal of this resting phase is to prepare for maintenance treatment with olaparib.

#### Maintenance treatment

The maintenance treatment phase starts after the level of Lymphocytes is back at the normal level defined with the threshold which is set to $1 \cdot 10^{10}$ cells. This threshold is the minimal level of Lymphocytes in the adult female body below which leukopenia is observed. After the treatment-resting phase, olaparib is administered to the virtual patient every 12 hours (twice daily) with a total dose per day equal to 300 mg as described in Text S4. The maintenance treatment with olaparib is administered to virtual ovarian cancer patients, until relapse or profound toxicity is observed, with the time

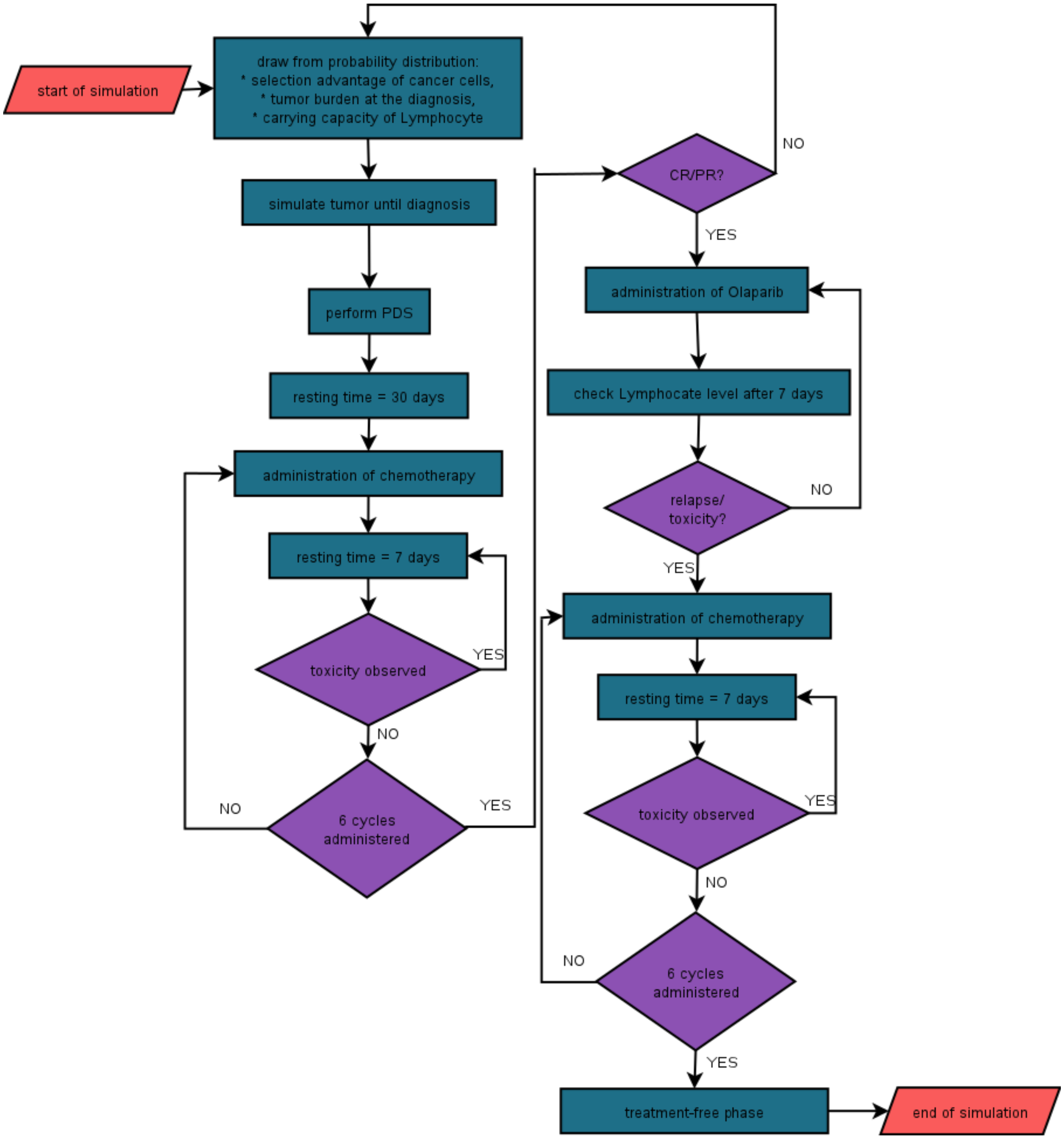


Figure 9: Schematic of developed simulator of advanced high-grade serous ovarian cancer.

limit of 2 years. After the first relapse, PFS is calculated as a time interval between time of patients randomization until relapse.

#### Second-line treatment

Next, the second-line treatment is administered in form of six cycles of platinum-based chemotherapy. The dosing of chemotherapy is the same as during the first-line treatment. This phase of simulation is incorporate into the mathematical model simulator to accurately compute the second progression-free survival.

#### Treatment-free phase

The last phase of simulation is a free (without drugs perturbation) tumor growth. This phase reflect the time when patient is in remission or (if the relapse is during the second-line treatment) patient is treated with palliative intent. Next, the second progression-free survival is calculated as a time interval between start of second-line treatment and the second relapse.

#### Drug toxicity management

Not all patient enrolled to SOLO-1 clinical trial and developed toxicity to olaparib has olaparib discontinued. During this trial, three methods of olaparib toxicity management were applied: discontinuation, dose reduction, and dose interruption. In the simulation, when toxicity is observed, a random number is sampled from multinomal distribution that has three outcomes (dose reduction, dose discontinuation, dose interruption). The probability that any of this method is selected is given with percentages available at Table S2.

In reality, some patients receive two or even three methods of adverse events management. However, it is not possible to extract those information from the available data about SOLO-1 clinical trial. So, in the model, we assume that only one method of adverse events management is applied.

Dose reduction is performed as follows. Firstly, the daily dose is reduced from 600 mg to 500 mg. If toxicity is still observed, the dosing is reduced again to 400 mg, next to 300 mg, and lastly to 200 mg. If still the last reduction does not help, the maintenance treatment with olaparib is stopped.

Dose discontinuation is simply stopping the treatment with no possibility to go back to the treatment again. This toxicity management is applied only when dose reduction or dose interruption will not help to ease the side effects.

Dose interruption is stopping the treatment for several weeks to allow white blood cells to go back to normal level. In our case, we investigate dose interruption from one to ten weeks. It is the most common method of toxicity management.

### Text S9. Computer simulator

The model is simulated as a branching process modeling following J. Reiter et al. 2013 [11]. The model is a discrete–state and discrete-time model where the system is updated at a constant time interval. Here, the system state is updated every twelve hours.

The model is implemented in MATLAB 2021 environment. Also, the lightspeed toolbox for efficient sampling from multinomial distribution is applied (https://github.com/tminka/lightspeed). As the model is not spacial (we assume a well-mixed system) and we update the system at every fixed time interval, the model implementation is efficient computationally.

#### Initial conditions

The simulations start from the number of Lymphocytes at carrying capacity level $K_{WBC}$ which is drawn from the uniform distribution as explained in Text S5, and a single sensitive cell.

#### Tumor cells

Tumor cells are growing exponentially with the following division:

$$b = \frac{1}{2} \cdot (1 + s) \tag{S14}$$

and death rate:

$$d = 1 - b. \tag{S15}$$

At time $(t + 1)$, $Y_1$ cells divide to an identical daughter cell, $Y_2$ cells divide with an additional resistance (to pt-based chemotherapy, olaparib, or both), and $Y_3$ cells die. Thus, the number of cancer cells at the next time step is sampled from multinomial probability distribution:

$$Prob([Y_1, Y_2, Y_3) = (y_1, y_2, y_3)] = \frac{N_j(t)!}{y_1!y_2!y_3!}[b_j(1-u)^{y_1} \cdot (b_j \cdot u)^{y_2} \cdot d^{y_3}] \tag{S16}$$

where $u = u_{pt-based\ chemotherapy} + u_{PARPi} + u_{cross} + u_{reverse}$, $X = y_1 + y_2 + y_3$, and $X_j(t + \Delta t) = X(t) + Y_1 - Y_3$.

We know, that $Y_2$ cells give rise to new subclone. Thus, if $Y_2 > 0$ in the simulation, then from multinomial distribution we can estimate the number of cells which give rise to a given subclone:

$$Prob[(Z_1, Z_2, Z_3, Z_4) = (z_1, z_2, z_3, z_4)] = \frac{M(t)!}{z_1!z_2!z_3!z_4!}[b_j \cdot u^{y_1}_{pt-based\ chemotherapy} \cdot b_j \cdot u^{y_2}_{PARPi} \cdot b_j \cdot u^{y_3}_{cross} \cdot b_j \cdot u^{y_4}_{reverse}] \tag{S17}$$

where $Z_1, Z_2$, $Z_3$, and $Z_4$ are number of cells which gain resistance to pt-based chemotherapy, olaparib, both or reverse of BRCA1/2 mutation respectively.

#### Lymphocytes

Lymhpocytes can undergo one of two processes: cell division or cell death. The division rate equals: $b = \frac{1}{2}(1 + s_{wbc}(t))$, where $s_{wbc}(t)$ is selection advantage. The death rate equals to $d = 1 - b$. From equations on $b$, we can notice that when $s_{WBC} = 0$, $d = b$ leading to the same probability of cell division and death. When $s_{WBC} = 1$, the model simplifies to the pure birth model where $d = 0$ and only division can occur.

We model the growth of Lymphocytes according to logistic growth model and thus:

$$s = s_{WBC} \cdot \left(1 - \frac{WBC}{K_{WBC}}\right), \tag{S18}$$

where $K_{WBC}$ is carrying capacity, $WBC$ is the current number of Lymphocytes and $s_{WBC}$ is maximal selection advantage. At each time step (generation), the number of Lymphocytes $WBC$ is sampled from binomial probability distribution:

$$Prob(WBC_1, WBC_2) = \frac{WBC(t)!}{wbc_1!wbc_2!}[b_j^{wbc_1} \cdot d^{wbc_2}] \tag{S19}$$

where $WBC(t) = wbc_1 + wbc_2$, and

$$WBC(t+1) = WBC(t) + wbc_1 - wbc_2. \tag{S20}$$

At time $(t+1)$, $WBC_1$ Lymphocytes divides and $WBC_2$ Lymphocytes die.

#### Pharmacokinetics

The drug concentration over time in the model is given with algebraic equation S3 (for pt-based chemotherapy) and S5 (for olaparib). As cispaltin is administrated by injection and olaparib by rapidly absorbed pills and both of these delivery methods are significantly faster than assumed simulation step (12h), we assume that, right after administration, the drug concentration increases by $C_{max}$. Thus, when the drug is administrated at time $t+1$, drug concentration changes in step-like manner according to the following equation:

$$C_{drug}(t+1) = C_{max_{drug}} + C_{drug}(t) \cdot exp(-k_{drug} \cdot t). \tag{S21}$$

### Text S10. Features extracted from virtual SOLO-1 clinical trial simulations

#### The list of features

Using the results from SOLO-1 clinical trial simulation, 12 features were extracted. All of them are listed in Table S6. Those features could be extracted from real patients and potentially be applied for optimization of maintenance treatment with olaparib.

Table S6: Features extracted from virtual SOLO-1 clinical trial

| Name | Description |
|---|---|
| Fitness | Fully-sensitive cancer cells selection advantage in percentage |
| Cx response | Response to chemotherapy, the fraction by which tumor is reduced |
| Selection under olaparib | Fully-sensitive cancer cells selection advantage during olaparib treatment |
| WBC naive | WBC level at diagnosis |
| WBC nadir | The lowest WBC level |
| Toxicity to PARPi | Are adverse events observed? |
| Subclones dx | Number of distinct subclones at the diagnosis |
| Subclones cx | Number of distinct subclones after chemotherapy |
| Fraction dx | Fraction of cancer cells resistant to chemotherapy and olaparib at the diagnosis |
| Residual tumor | Tumor size after surgery |
| $M_{diagnosis}$ | Tumor burden at the diagnosis |
| Maintenance time | Duration of maintenance treatment |

#### Univariate Cox regression

Using extracted features and survival data (the first PFS), univariate Cox regression was performed for identification of the first PFS biomarkers. The goal is to identify which extracted features could alone be a good biomarker of response to olaparib maintenance treatment.

Cox regression was performed in the R environment using survival package. 10-fold cross-validation was performed using results from 10,000 virtual patients simulations. The model performance was assessed using the Harell concordance index.

### References


[1] Average Weight for Women: By Age, By Height, Tables, and More.

[2] Ovarian Cancer Statistics — Ovarian Cancer Research Alliance.

[3] A. H. Calvert, D. R. Newell, L. A. Gumbrell, S. O'Reilly, M. Burnell, F. E. Boxall, Z. H. Siddik, I. R. Judson, M. E. Gore, and E. Wiltshaw. Carboplatin dosage: prospective evaluation of a simple formula based on renal function. *Journal of Clinical Oncology*, 7(11):1748–1756, 1989.

[4] Susanna L. Cooke and James D. Brenton. Evolution of platinum resistance in high-grade serous ovarian cancer, nov 2011.

[5] Rachel Karchin, Tibor Antal, Dewey Kim, Martin A Nowak, Bert Vogelstein, Hannah Carter, Sining Chen, Ivana Bozic, Kenneth W Kinzler, and Hisashi Ohtsuki. Accumulation of driver and passenger mutations during tumor progression. *Proceedings of the National Academy of Sciences*, 107(43):18545–18550, oct 2010.

[6] Emilia Kozłowska, Anniina Färkkilä, Tuulia Vallius, Olli Carpén, Jukka Kemppainen, Seija Grénman, Rainer Lehtonen, Johanna Hynninen, Sakari Hietanen, and Sampsa Hautaniemi. Mathematical Modeling Predicts Response to Chemotherapy and Drug Combinations in Ovarian Cancer. *Cancer research*, 78(14):4036–4044, jul 2018.

[7] Ashley M. Laughney, Sergi Elizalde, Giulio Genovese, and Samuel F. Bakhoum. Dynamics of Tumor Heterogeneity Derived from Clonal Karyotypic Evolution. *Cell Reports*, 12(5):809–820, aug 2015.

[8] Behzad Mansoori, Ali Mohammadi, Sadaf Davudian, Solmaz Shirjang, and Behzad Baradaran. The different mechanisms of cancer drug resistance: A brief review, sep 2017.

[9] Tariq Masoodi, Sarah Siraj, Abdul K. Siraj, Saud Azam, Zeeshan Qadri, Sandeep K. Parvathareddy, Asma Tulbah, Fouad Al-Dayel, Hamed AlHusaini, Osama AlOmar, Ismail A. Al-Badawi, Fowzan S. Alkuraya, and Khawla S. Al-Kuraya. Genetic heterogeneity and evolutionary history of high-grade ovarian carcinoma and matched distant metastases. *British Journal of Cancer*, 122(8):1219–1230, apr 2020.

[10] Kathleen Moore, Nicoletta Colombo, Giovanni Scambia, Byoung Gie Kim, Ana Oaknin, Michael Friedlander, Alla Lisyanskaya, Anne Floquet, Alexandra Leary, Gabe S. Sonke, Charlie Gourley, Susana Banerjee, Amit Oza, Antonio González-Martín, Carol Aghajanian, William Bradley, Cara Mathews, Joyce Liu, Elizabeth S. Lowe, Ralph Bloomfield, and Paul Disilvestro. Maintenance Olaparib in Patients With Newly Diagnosed Advanced Ovarian Cancer, 2019.

[11] Johannes G. Reiter, Ivana Bozic, Krishnendu Chatterjee, and Martin A. Nowak. TTP: Tool for Tumor Progression. In *Lecture Notes in Computer Science (including subseries Lecture Notes in Artificial Intelligence and Lecture Notes in Bioinformatics)*, volume 8044 LNCS, pages 101–106. 2013.

[12] J M Riedinger, B Coudert, I Barillot, G Buffenoir, F Mayer, P Fargeot, J Cuisenier, J Guerrin, Riedinger J.-M., Coudert B., Barillot I., Buffenoir G., Mayer F., Fargeot P., Cuisenier J., Guerrin J., and Riedinger J.M. Clinical value of the estimation of growth kinetics of primary ovarian cancer recurrences by CA125 doubling time. *Bulletin du cancer*, 84(9):855–860, 1997.

[13] John O Schorge, Christopher McCann, and Marcela G Del Carmen. Surgical debulking of ovarian cancer: what difference does it make? *Reviews in obstetrics and gynecology*, 3(3):111–7, 2010.

[14] Mayo Clinic Staff. Creatinine test Results - Mayo Clinic, 2013.

[15] Kaiming Sun, Keith Mikule, Zebin Wang, Grace Poon, Aparajitha Vaidyanathan, Gillian Smith, Zhi Yi Zhang, Jeffrey Hanke, Sridhar Ramaswamy, and Jing Wang. A comparative pharmacokinetic study of PARP inhibitors demonstrates favorable properties for niraparib efficacy in preclinical tumor models. *Oncotarget*, 9(98):37080–37096, dec 2018.

[16] B. Tummers. DataThief III, 2006.

[17] Marc J Williams, Benjamin Werner, Chris P Barnes, and Trevor A Graham. Identification of neutral tumor evolution across cancer types. *Nature Genetics*, 48(3):238–244, 2016.

[18] Ritsu Yamamoto, Shinichiro Minobe, Masanori Kaneuchi, Noriaki Sakuragi, Seiichiro Fujimoto, Yoshiaki Ishizaki, Hiroka Domon, Hitoshi Hareyama, Chikara Sato, Takahumi Fujino, Isao Kawaguchi, Tatsumi Yamaguchi, Toshio Fujimoto, and Ken Yoshiaki. A phase I/II study of carboplatin and paclitaxel in patients with epithelial ovarian cancer. *Japanese Journal of Clinical Oncology*, 32(4):128–134, apr 2002.
